# Dual-systems models of the genetic architecture of impulsive personality traits: Neurogenetic evidence of distinct but related factors

**DOI:** 10.1101/2023.02.10.23285725

**Authors:** Alex P. Miller, Ian R. Gizer

## Abstract

**Background:** Dual-systems models provide a parsimonious framework for understanding the interplay between cortical and subcortical brain regions relevant to impulsive personality traits (IPTs) and their associations with psychiatric disorders. Despite recent developments in multivariate analysis of genome-wide association studies (GWAS), molecular genetic investigations of these models have not been conducted.

**Methods:** Using extant IPT GWAS, we conducted confirmatory genomic structural equation models (GenomicSEM) to empirically evaluate dual-systems models of the genetic architecture of IPTs. Genetic correlations between results of multivariate GWAS of dual-systems factors and GWAS of relevant cortical and subcortical neuroimaging phenotypes (regional/structural volume, cortical surface area, cortical thickness) were calculated and compared.

**Results:** Evaluation of GenomicSEM model fit indices for dual-systems models suggested that these models highlight important sources of shared and unique genetic variance between top-down and bottom-up constructs. Specifically, a dual-systems genomic model consisting of sensation seeking and lack of self-control factors demonstrated distinct but related sources of genetic influences (*r*_*g*_=.60). Genetic correlation analyses provided evidence of differential associations between dual-systems factors and cortical neuroimaging phenotypes (e.g., lack of self-control negatively associated with cortical thickness, sensation seeking positively associated with cortical surface area). However, no significant associations were observed for subcortical phenotypes inconsistent with hypothesized functional localization of dual-systems constructs.

**Conclusions:** Dual-systems models of the genetic architecture of IPTs tested here demonstrate evidence of shared and unique genetic influences and associations with relevant neuroimaging phenotypes. These findings emphasize potential advantages in utilizing dual-systems models to study genetic influences for IPTs and transdiagnostic associations with psychiatric disorders.

---

Research has shown that trait impulsivity and disinhibition (i.e., impulsive personality traits [IPTs]) confer transdiagnostic risk for a multitude of psychiatric disorders with a particularly important role in disorders of the externalizing spectrum (Beauchaine, Zisner, & Sauder, 2017; Creswell, Wright, Flory, Skrzynski, & Manuck, 2019; S. Johnson, Carver, & Joormann, 2013). Broadly, IPTs are characterized by a lack of self-control and forethought of behavioral consequences in response to more temporally salient external stimuli or internal impulses (Whiteside & Lynam, 2001). Twin studies have demonstrated that IPTs are significantly heritable, though heritability estimates can vary substantially, in part, based on the specific facet under study (Bezdjian, Baker, & Tuvblad, 2011; Friedman et al., 2020; Gustavson et al., 2019). Similarly, neuroimaging studies suggest some heterogeneity in findings across neural correlates of IPTs, but also support a general pattern of differential brain morphology associated with cortical regions involved in cognitive control and attention, subcortical regions involved in reward and emotion-processing, and connections between them (e.g., mesocorticolimbic and frontostriatal pathways; Pan et al., 2021; S. Johnson, Elliott, & Carver, 2020).

In aggregate, genetic and neurobiological studies of IPTs have led to the hypothesis that IPTs and related brain morphology may serve as useful endophenotypes for the study of psychiatric disorders, especially those of the externalizing spectrum (e.g., substance use disorders [SUDs]; Congdon & Canli, 2005; Jonas & Markon, 2014; Cyders, Coskunpinar, & VanderVeen, 2016; Verdejo-Garcia & Albein-Urios, 2021). The endophenotype concept argues that studying genetic influences on relevant neuropsychological traits (e.g., IPTs) underlying risk for a manifest disorder (e.g., SUDs) may help to identify shared functional neurobiological and genetic factors (Hall & Smoller, 2010; Kendler & Neale, 2010). Despite this promise, conceptualizations of IPTs exhibit substantial heterogeneity and though this variability may be meaningful, it may also contribute to a lack of clarity about the mechanisms underlying the relation between IPTs and clinical presentation (Strickland & M. Johnson, 2021). Thus, the use of overly multidimensional IPTs as endophenotypes in genetic studies may hamper the ability to detect important and biologically relevant shared causal loci. To increase power to detect shared neurogenetic etiology, the current study relies on an established IPT model rooted in developmental and neurobiological frameworks: the dual-systems model.

The dual-systems model provides a parsimonious framework for understanding the interplay between cortical and subcortical brain regions relevant to impulsive behavior. Specifically, the model posits that impulsive behaviors are the result of two complementary neurobiological systems associated with distinct neural substrates acting in dynamic tension to influence behavior: (1) a bottom-up system, involving activation of subcortical regions involved in reward (e.g., sensation seeking) and/or emotion-based drive (e.g., urgency), and (2) a top-down system, involving activation of prefrontal cortical regions involved in effortful control and forethought (e.g., self-control; Casey, Getz, & Galván, 2008; Carver & S. Johnson, 2018; Dalley & Robbins, 2017; Ernst, 2014; Shulman et al., 2016; Steinberg et al., 2008).

Notably, the dual-systems model is not only compatible with empirical neurocognitive observations but also with developmental theory (Steinberg et al., 2018). The transition from adolescence to adulthood is characterized by a developmental ‘spike’ in risky, impulsive behaviors driven by rapid increases in sensitivity to reward and affective salience (e.g., sensation seeking, urgency) paired with relatively slower maturation of cortical structures that govern inhibition, planning, and self-control (Lopez-Vergara, Spillane, Merrill, & Jackson, 2016; Shulman, Harden, Chein, & Steinberg, 2016). While being especially relevant in youth, the dual-systems model is also appropriate for explaining impulsive behaviors across the lifespan (Duell et al., 2016).

Molecular genetic investigations of dual-systems models could help to define more homogenous sub-groups of individuals if genetic liability for psychiatric disorders can be parsed into risk for increased bottom-up approach behaviors or lack of top-down cognitive control. Prior twin studies examining genetic influences for distinct top-down and bottom-up IPTs have indicated that observed overlap between the two systems may be due to shared genetic factors (Ellingson, Verges, Littlefield, Martin, & Slutkse, 2013; Ellingson et al., 2018; Hur & Bouchard, 1997). Thus, further study of the shared genetic influences between IPTs and the underlying neurobiology of these traits is needed. Recent advances in modeling of post-genome-wide association study (GWAS) summary statistics allows for more specific examination of the interrelations among genetic influences for these traits.

The aims of the present study were two-fold. First, the study aimed to leverage extant IPT GWAS summary statistics and an advanced multivariate GWAS approach (i.e., GenomicSEM; Grotzinger et al., 2019) to model important sources of unique and shared variance associated with dual-systems constructs. Separable factors of two distinct bottom-up constructs and a single top-down construct were hypothesized and empirically evaluated. These factors map onto reward-based bottom-up tendencies (sensation seeking), emotion-based bottom-up tendencies (urgency), and a common top-down factor (lack of self-control). Second, the study aimed to distinguish between neurogenetic influences related to top-down and bottom-up constructs by examining genetic correlations with neuroimaging phenotypes. While the post-GWAS genetic correlation analyses of this study were comparatively exploratory in nature, it was expected that dual-systems constructs would exhibit separable but overlapping genetic associations with brain regions implicated by previous research and theory (e.g., lack of self-control related to cortical phenotypes, sensation seeking and urgency related to subcortical phenotypes).

## Methods

Descriptions of GWAS summary statistics for all IPT and neuroimaging phenotypes used in this study are included in Table 1. Summary statistics were restricted to individuals of European ancestry and common variants (minor allele frequency [MAF] > 0.01) for the present analyses. See Supplementary Methods for descriptions of genotyping, imputation, quality control, meta-analytic procedures, and additional measurement information for discovery GWAS.

**Table 1.**
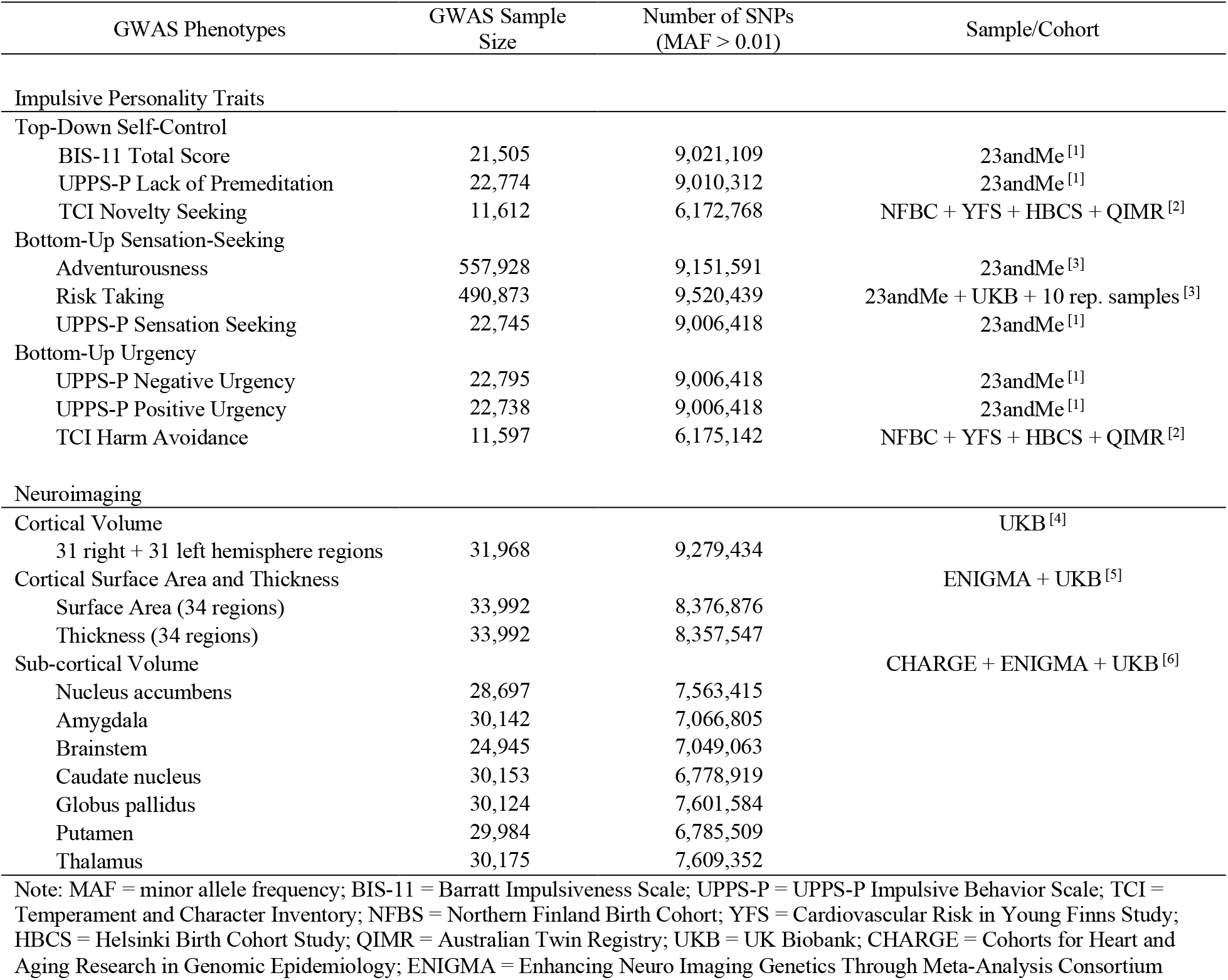
Overview of GWAS used in study.

### Impulsive Personality Trait GWAS

GWAS IPT phenotypes were primarily measured using the brief version of the UPPS-P measuring five subscales: lack of premeditation, lack of perseverance, negative urgency, positive urgency, and sensation seeking (Cyders, Littlefield, Coffey, & Karyadi, 2014), the BIS-11 measuring various aspects of impulsive personality (Patton, Stanford, & Barratt, 1995), and Cloninger’s Temperament and Character Inventory (TCI) measuring four primary domains of temperament: novelty seeking, harm avoidance, reward dependence, and persistence (Cloninger, Przybeck, Svrakic, & Wetzel, 1994). GWAS summary statistics for IPT phenotypes were obtained from three primary sources: the UK Biobank (UKB; risk taking, Linnér et al., 2019), direct-to-consumer genetics company 23andMe, Inc. (Sunnyvale, CA; BIS-11, UPPS-P, risk taking, and adventurousness; Linnér et al., 2019; Sanchez-Roige et al., 2019), and a meta-analytic sample comprised of four European ancestry cohorts (TCI harm avoidance and novelty seeking; Service et al., 2012).

*A priori* hypotheses regarding appropriate phenotype structure for dual-systems models, given available GWAS data and prior phenotypic research, led to specification of two separate two-factor confirmatory genomic structural models. The first model, hereafter referred to as the urgency-self-control (UGSC) model, consisted of a bottom-up ‘urgency’ factor indexing genetic influences for high urgency, i.e., emotion-based rash action, and a top-down ‘(lack of) self-control’ factor indexing genetic influences for low self-control, lack of planning, and lack of forethought (see Figure 1A). The second model, hereafter referred to as the sensation seeking-self-control (SSSC) model, consisted of a bottom-up ‘sensation seeking’ factor indexing genetic influences for high sensation seeking, i.e., reward-based rash action, and the same top-down ‘(lack of) self-control’ factor (see Figure 1B). Thus, each model is characterized by the presence of the same ‘top-down’ (lack of) self-control genetic factor and a unique ‘bottom-up’ genetic factor conferring risk for either reward-based (sensation seeking) or emotion-based (urgency) drive behaviors hypothesized to act in dynamic tension with the ‘top-down’ genetic factor. Notably, models containing all three factors (i.e., correlated three-factor model) failed to converge. GWAS indicator selection and rationale are described below for each factor.

**Figure 1.**
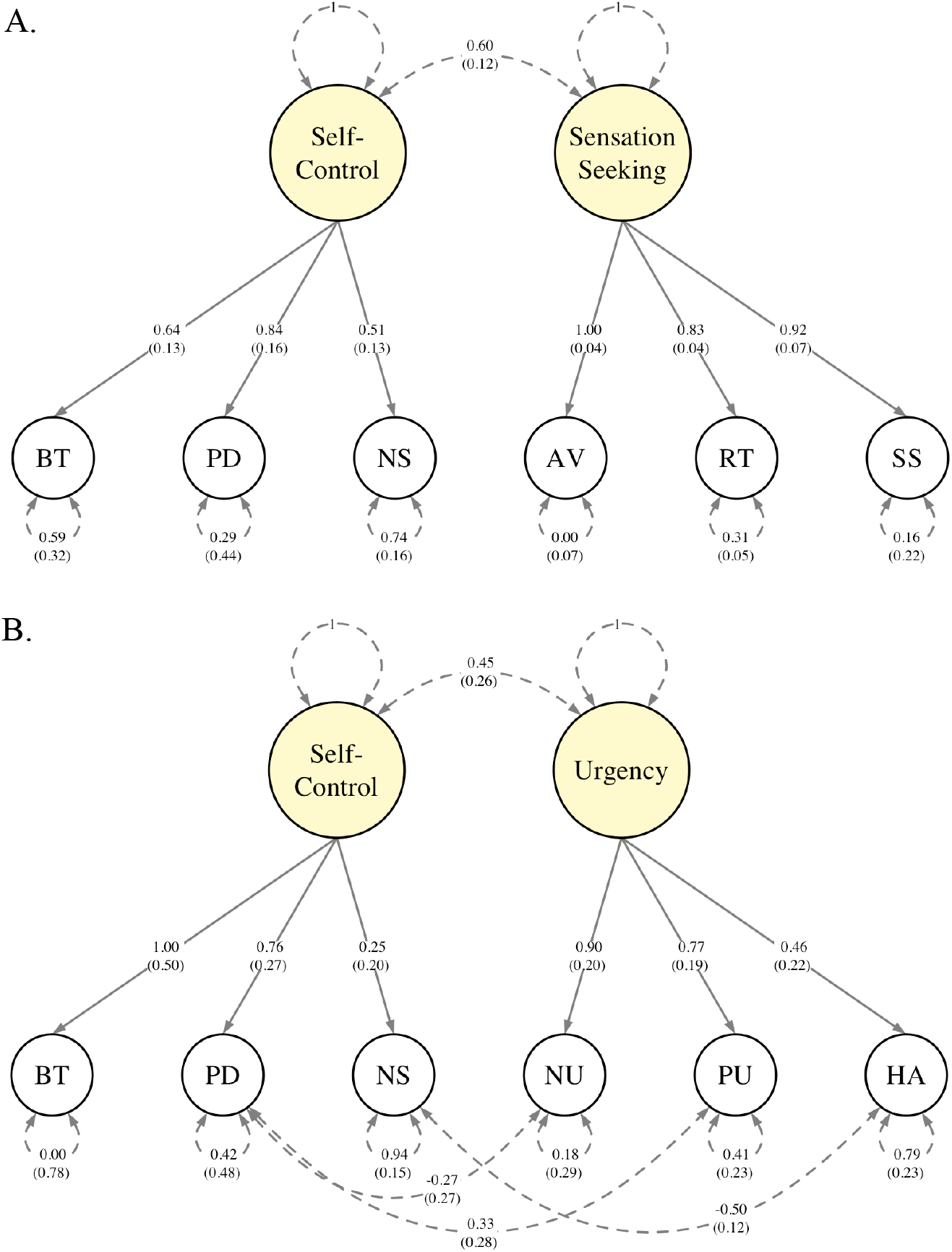
Final path diagram of the SSSC (A) and UGSC (B) dual-systems models estimated with GenomicSEM. Presented parameters are standardized and *SE* are shown in paratheses. Variances and covariances shown as dashed lines and factor loadings shown as solid lines. Note, “self-control” denotes (lack of) self-control factor. See Supplementary Tables 3 and 4 for model fit indices. BT = BIS-11 Total Score; PD = UPPS-P Lack of Premeditation; NS = TCI Novelty Seeking; AV = Adventurousness; RT = Risk Taking; SS = UPPS-P Sensation Seeking; NU = UPPS-P Negative Urgency; PU = UPPS-P Positive Urgency; HA = TCI Harm Avoidance

### Lack of Self-Control

GWAS of the BIS-11 total score, the UPPS-P lack of premeditation subscale, and the TCI novelty seeking scale were specified as indicators of the top-down (lack of) self-control factor. While some literature reviews have clustered novelty seeking with sensation seeking measures (e.g., Fischer, G. Smith, & Cyders, 2008; Stautz & Cooper, 2013), empirical evidence across a number of samples, including the UPPS development sample (Whiteside & Lynam, 2001), suggests that novelty seeking is strongly associated with lack of premeditation and planning and only weakly associated with sensation seeking (C. Evren, Durkaya, B. Evren, Dalbudak, & Cetin, 2012; Savvidou et al., 2017; Vonmoos et al., 2013).

Prior research has also suggested that BIS-11 subscales demonstrate poor psychometric properties (Morean et al., 2014; Reise, Moore, Sabb, Brown, & London, 2013), and that BIS-11 total scores and UPPS-P lack of premeditation subscale scores exhibit substantial genetic and phenotypic overlap related to lack of self-control and planning prior to action (Gustavson et al., 2020; Sanchez-Roige et al., 2019).

### Urgency

GWAS of the UPPS-P negative and positive urgency subscales and the TCI harm avoidance scale were specified as hypothesized indicators of the bottom-up urgency factor. Empirical studies have suggested that negative and positive urgency are highly correlated and together may represent a common transdiagnostic risk factor for psychiatric disorders (Billieux et al., 2021). This notion is substantiated by high phenotypic and genetic correlations between negative and positive urgency (*r*=.59; *r*_*g*_*=*.74) in the 23andMe sample (Sanchez-Roige et al., 2019). While the TCI harm avoidance scale is not strictly considered to be a measure of IPT, it is thought to reflect a tendency to respond intensely to aversive stimuli and negative affect with loss of control of behavioral responses (Cloninger, 1987) and has been shown to be moderately correlated with negative urgency in clinical samples (*r*=.28-.55; Jiménez-Murcia et al., 2020; Savvidou et al., 2017).

### Sensation Seeking

A GWAS of risk taking from a meta-analysis including both UKB and 23andMe samples (see Supplementary Methods for GWAS meta-analysis description and Supplementary Figure 1 for quantile-quantile [Q-Q] plot) was specified along with a GWAS of a Likert-scale item assessing adventurousness (“Would you consider yourself to be more cautious or more adventurous?”; Linnér et al., 2019) and the UPPS-P sensation seeking subscale as indicators of the bottom-up sensation seeking factor.

### Neuroimaging GWAS

Three primary sets of neuroimaging GWAS were utilized for genetic correlation analyses (see Table 1 and Supplementary Methods). The first set included UKB GWAS of 62 (31 left/right hemisphere) cortical parcellation phenotypes generated via the Desikan-Killiany-Tourville atlas (Klein & Tourville, 2012) and obtained from the Oxford Brain Imaging Genetics web server (S. Smith et al., 2021). The second set included GWAS of 34 cortical surface area (SA) and thickness (TH) parcellation phenotypes (Grasby et al., 2020) generated via the Desikan-Killiany atlas (Desikan et al., 2006). The third set included GWAS of seven subcortical structures (Satizabal et al., 2019).

### Data Analysis

As GWAS data for TCI phenotypes (novelty seeking and harm avoidance) contained summary statistics for only ∼1.2 million variants, which is inadequate for multivariate modeling of GWAS using GenomicSEM, summary statistics imputation was implemented using *ssimp* (v0.5.6; Rüeger, McDaid, & Kutalik, 2018). Imputation and data analytics procedures are described in more depth in Supplementary Methods.

### Genomic Factor Models

GenomicSEM (version 0.0.5; Grotzinger et al., 2019), an extension of linkage disequilibrium score regression (LDSC; Bulik-Sullivan et al., 2015), was employed using diagonally weighted least squares estimation and unit variance identification (i.e., variance of each latent factor is fixed to 1) to conduct confirmatory factor analyses empirically modeling the genetic associations among top-down and bottom-up phenotypes. Two primary models were tested at this stage: (1) the SSSC model reflecting shared genetic architecture between top-down lack of self-control and bottom-up sensation seeking, and (2) the UGSC model reflecting shared genetic architecture between top-down lack of self-control and bottom-up urgency. Model fit was assessed using *χ*^2^ tests, the comparative fit index (CFI; Bentler, 1990), the standardized root mean square residual (SRMR; Bentler, 1995), and the Akaike information criterion (AIC; Akaike, 1974). Additionally, given that dual-systems models contained indicator GWAS from the same measure (i.e., UPPS-P) and sample (i.e., 23andMe) across correlated latent factors, follow-up models allowing within-measure cross-factor residuals to covary were fit with changes in model fit assessed using *χ*^2^ difference tests (Δ*χ*^2^).

Single latent factor models were also specified separately for each of the three dual-systems constructs in order to: (1) minimize the effect of uneven sample sizes between traits as described in Supplementary Methods, (2) increase the number of variants tested for each trait as variants are excluded using listwise deletion across indicators, and (3) limit any potential estimation bias introduced by residual covariance structures as described above. Because these single latent factor models each had three indicator GWAS (i.e., fully saturated just-identified models with zero degrees of freedom), model fit indices were unavailable, and fit was instead assessed by examining the significance of indicator loadings and residual variances.

### Multivariate GWAS

Following examination of genomic structural models, multivariate GWAS were conducted to estimate SNP associations with each latent dual-systems genetic factor using GenomicSEM. Individual SNP effects were estimated for each latent genetic factor if they were available across all indicator GWAS, had a MAF ≥ 0.5%, and were present in the 1000 Genomes Project Phase 3 v5 reference panel (The 1000 Genomes Project Consortium, 2015). Effective sample sizes for each latent factor (*N*_*eff*_) were estimated using the approach described by Mallard, Linnér, et al. (2022). SNP-based heritability estimates 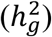 of latent genetic factors derived using univariate LDSC and *N*_*eff*_ are more accurately referred to as genetic variances (Mallard, Linnér, et al., 2022) and are subsequently denoted by *ζ*_*g*_ to differentiate between these two indices. To identify SNP effects not fully mediated by the specified single latent factor (i.e., common pathway model), follow-up multivariate GWAS including unique pathways were conducted to calculate *Q*_SNP_ tests of heterogeneity. SNPs with genome-wide significant (GWS; *P*<5×10^−8^) *Q*_SNP_ statistics exert effects on genetic indicators independent of the latent factor (Grotzinger et al., 2019). Thus, these SNPs were removed from model-derived GWAS summary statistics to reduce heterogeneity in the latent genetic factors for downstream analyses.

### Neuroimaging Genetic Correlation Analyses

LDSC genetic correlation analyses were conducted to examine whether dual-systems constructs differed with respect to their genetic overlap with regional cortical volume, SA, TH, and subcortical structural volume. GenomicSEM was used to calculate genetic correlations between each genetic factor and each neuroimaging phenotype with a 5% FDR correction used to account for multiple testing within each imaging phenotype set for each latent construct separately. To determine whether correlations with each neuroimaging phenotype differed between paired top-down and bottom-up constructs, *χ*^2^ tests were used to evaluate the null hypothesis that each pair of genetic correlations could be constrained to equality (Demange et al., 2021).

## Results

### Genomic Factor Models

Preliminary univariate and bivariate LDSC estimates for all indicator GWAS are shown in Supplementary Tables 1-2 and Supplementary Figure 2. SNP-based heritability estimates were all significant at 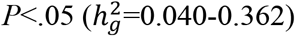. Ratio values, (LDSC intercept−1)/(mean *χ*^2^−1), which provide an estimate of the proportion of polygenic signal due to biased inflation, were not significantly different from zero for most traits, suggesting negligible inflation of test statistics from sources other than true genetic effects (e.g., uncontrolled population stratification). Of note, because the TCI novelty seeking and harm avoidance GWAS were likely underpowered, as evidenced by *λ*_GC_, mean *χ*^2^, and LDSC intercept values below 1, ratio values were undefined, and 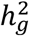 estimates (0.362-0.305) are likely inflated. Nevertheless, LDSC genetic correlations were consistent with hypothesized predictions as they demonstrated appreciable clustering among factor indicator GWAS for each dual-systems genetic latent factor (Supplementary Figure 2). Further, TCI GWAS contributed to the polygenic signal and heritability of subsequent latent genetic factors as described in the *Multivariate GWAS* section below.

**Figure 2.**
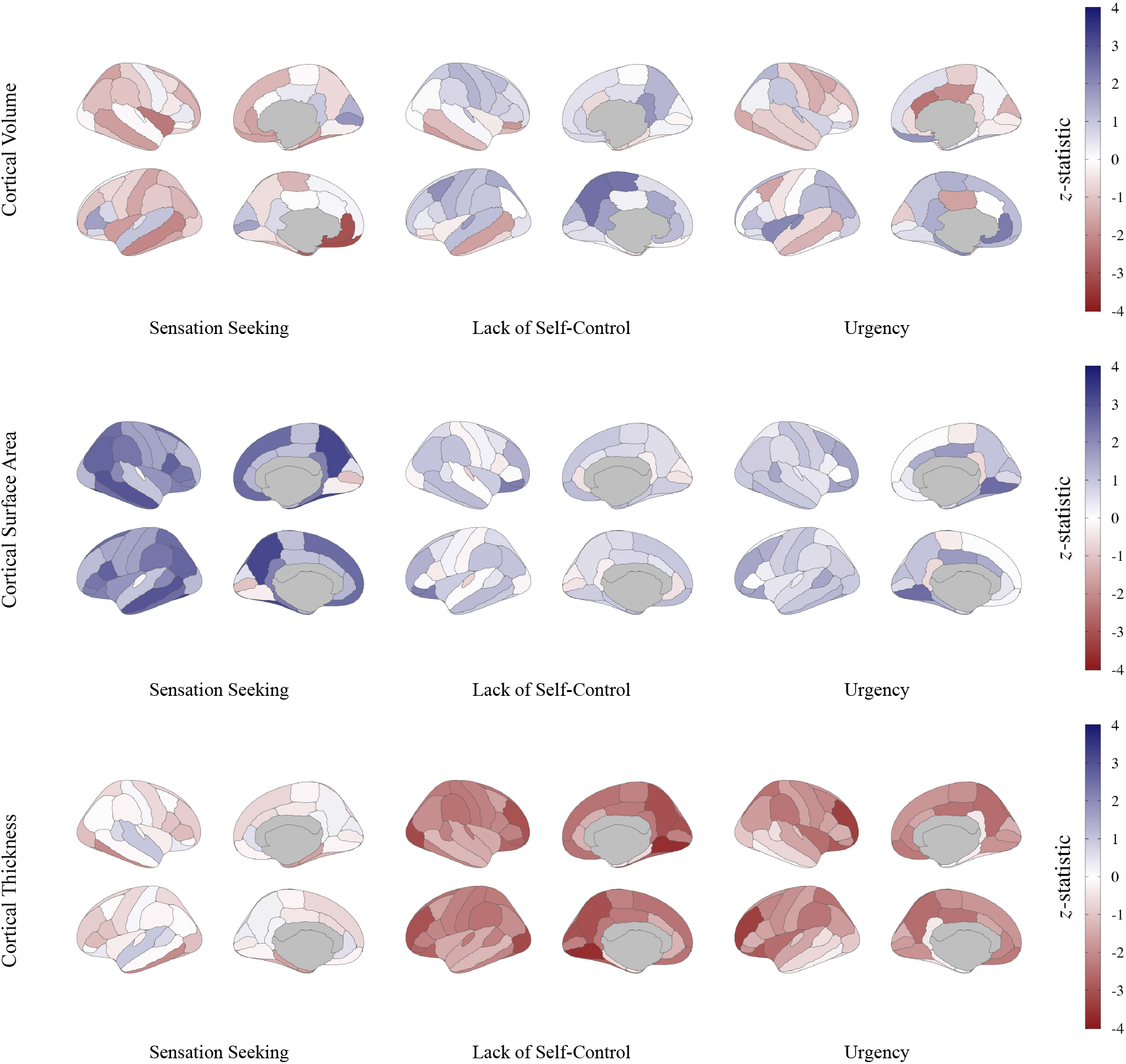
Genetic correlations between dual-systems factors and regional cortical brain volume, cortical surface area, and cortical thickness. Cortical patterning of genetic correlations plotted as *z*-statistics (blue = positive correlation, red = negative correlation) across IPT dual-systems traits for cortical regional volume (top) according to the Desikan-Killiany-Tourville atlas (Klein and Tourville, 2012), and cortical regional surface area (middle) and thickness (bottom) according to the Desikan-Killiany atlas (Desikan et al., 2006). Plots were constructed using the *ggseg* package in R (Mowinckel & Vidal-Piñeiro, 2020).

Structural GenomicSEM analyses showed that the correlated factors dual-systems models provided good fit to the genetic covariance matrices. Specifically, the SSSC model provided good fit to the data (*χ*^2=^10.67, *df*=8, *P*=.22, AIC=36.67, CFI=1.00, SRMR=.09) that was not improved when within-measure cross-factor residuals of the UPPS-P sensation seeking and lack of premeditation indicator GWAS were allowed to covary (*χ*^2=^10.69, *df*=7, *P*=.15, AIC=38.69, CFI=1.00, SRMR=.08; Δ*χ*^2^=-0.02, *df*=1, *P*=.887), allowing the residual covariance path to be dropped from the final model. The bottom-up sensation seeking factor and the top-down (lack of) self-control factor in the SSSC model were strongly correlated (*r*_*g*=_.60, *SE=*0.12, *P*=2.15×10^−^ _7_; Figure 1A; Supplementary Table 3).

The initial UGSC model provided poor fit to the data (*χ*^2=^56.15, *df*=8, *P*=2.63×10^−9^, AIC=82.15, CFI=.56, SRMR=.16), but fit was drastically improved when paths allowing within-measure cross-factor residual covariances between the UPPS-P negative and positive urgency indicator GWAS and the UPPS-P lack of premeditation indicator GWAS and between the TCI harm avoidance and novelty seeking indicator GWAS were added (*χ*^2=^10.67, *df*=5, *P* =.058, AIC=42.67, CFI=.95, SRMR=0.09; Δ*χ*^2^=45.49, *df*=3, *P*=7.29×10^−10^). The bottom-up urgency factor and the top-down (lack of) self-control factor in the UGSC model exhibited a moderate, but nonsignificant, correlation (*r*_*g*=_.45, *SE=*0.23, *P*=.063; Figure 1B; Supplementary Table 4).

While model fit indices were not available for the single latent factor dual-systems models, factor loadings were acceptable to large (*λ*=0.38-1.00) and mostly significant at *P*<.05 with the exception of the TCI novelty seeking GWAS (*λ*=0.38, *SE*=0.22, *P*=.083). Similarly, residual variances were generally small and non-significant with the exception of the TCI novelty seeking and harm avoidance GWAS (*ε*_NS_=0.86, *SE*=0.19, *P=*6.83×10^−5^; *ε*_HA_=0.78, *SE*=0.24, *P=*.001, respectively) and the risk taking GWAS (*ε*_RT_=0.32, *SE*=0.05, *P=*1.52×10^−9^). For full output of model parameters and path diagrams for dual-systems single latent factor models, see Supplementary Table 5 and Supplementary Figure 3, respectively.

### Multivariate GWAS

The multivariate GWAS analysis of the sensation seeking factor identified 1092 independent GWS variants (mapped to 262 independent genomic loci; *N*_*eff*_=710,971), and no SNPs displayed significant heterogeneity (GWS *Q*_SNPs_) in individual effects for the sensation seeking factor. Although there is early lift-off in the Q-Q plot (Supplementary Figure 4), univariate LDSC analysis indicated that the results were not due to uncontrolled inflation, bias, or stratification (ratio value=0.01, *SE*=0.01), but rather reflect the extensive polygenicity of this trait (*ζ*_*g=*_0.087, *SE=*0.003; mean *χ*^2^=2.26; Supplementary Table 6). For more detailed description of sensation seeking GWAS results see Miller & Gizer (2023).

In contrast, no variants in the (lack of) self-control or urgency GWAS reached GWS. No GWS *Q*_SNPs_ were identified for the (lack of) self-control factor, but 323 GWS *Q*_SNPs_ were identified for the urgency factor. These were removed from the urgency factor summary statistics prior to effective sample size calculation and downstream analyses. Univariate LDSC analyses of the (lack of) self-control (*N*_*eff*_=27,656) and urgency (*N*_*eff*_=28,316) factors suggested that these traits display significant genetic variance ([lack of] self-control *ζ*_*g*_=0.072, *SE*=0.019; urgency *ζ*_*g*_=0.093, *SE*=0.022; Supplementary Table 6). However, examination of Q-Q plots (Supplementary Figures 5-6), lower observed mean *χ*^2^ values (1.03–1.06), and the absence of GWS associations imply that current sample sizes for these traits lack power necessary to identify meaningful variant-level loci.

### Genetic Correlations with Neuroimaging Phenotypes

Key findings from neuroimaging genetic correlation analyses include the following significant associations (*P*_FDR_<.05): (1) sensation seeking was positively correlated with cortical surface area (SA) across several regions, though correlations were small (.07<|*r*_*g*_|<.10); (2) (lack of) self-control was negatively correlated with cortical thickness (TH) across the majority of tested regions with moderate effect sizes (.22<|*r*_*g*_|<.44); and (3) urgency was negatively correlated with TH in the rostral middle frontal cortex (*r*_*g*=_-.39, *P*_FDR_=.031). Dual-systems factors were not associated with regional brain volumes nor subcortical structural volumes following FDR correction (Supplementary Table 7-8).

Observed differences in genetic correlations with SA and TH were generally robust across traits (Figure 4; Supplementary Tables 9-10). Broadly, genetic correlations between dual-systems factors and SA were in the positive direction while genetic correlations with TH were negative. However, (lack of) self-control and urgency were only nominally associated with SA in two and one regions, respectively (*P*=.012-.025). Sensation seeking, in contrast, was associated with SA following FDR correction across more than 25% of the regions tested (*P*_FDR_<.05) with equal representation in the frontal, parietal, and temporal lobes. Notably, *χ*^2^ tests constraining the magnitude of these correlations to equality across traits were generally non-significant (*P*_diff_>.05), suggesting that these associations were non-specific to sensation seeking. Conversely, (lack of) self-control was negatively correlated with TH across more than 60% of cortical regions tested (*P*_FDR_<.05) with the greatest representation in the frontal and parietal lobes, and correlations were significantly larger than those between sensation seeking and TH (*P*_diff_<.05 for ∼65% of regional comparisons). There were no significant correlations between TH and sensation seeking.

## Discussion

The current study represents the first investigation of the latent genetic structure of the dual-systems model of IPTs using molecular genetic data. Genomic factor analyses supported the distinct but related hypothesized genetic components of the tested dual-systems models. The SSSC model was strongly supported with fit indices demonstrating that the bottom-up sensation seeking factor and top-down (lack of) self-control factor represented separable, though correlated (*r*_*g*=_.60), constructs. These results are consistent with prior twin studies of the dual-systems model, which reported similar support for these constructs and a similar genetic correlation between them (Ellingson et al., 2013; Hur & Bouchard, 1997). After accounting for within-measure cross-factor residual covariances, the UGSC model was also supported, showing satisfactory model fit, and a modest, though nonsignificant correlation between the bottom-up urgency factor and the top-down (lack of) self-control factor (*r*_*g*=_.45). The lack of significance here likely resulted from divergent associations between the (lack of) self-control factor and the urgency factor indicators (Supplementary Figure 2), though the observed correlation is within the relatively wide range of previous estimates using similar traits from both twin and molecular genetic studies (e.g., *r*_*g*=_.26-.64; Gustavson et al., 2019, 2020). As noted, the present study is the first to investigate the latent genetic architecture of dual-systems models using molecular genetic data, and the first of any type to investigate the genetic architecture of an urgency and (lack of) self-control dual-systems model. Thus, while prior studies have examined genetic correlations amongst IPTs using molecular genetic approaches (e.g., Sanchez-Roige et al., 2019, 2022), investigations of the latent genetic architecture underlying dual-systems models more specifically are limited to a small number of behavior genetic studies (Ellingson et al., 2013; 2018; Harden et al., 2017), and as a result, the findings from the present study represent a novel extension of this prior work.

To further evaluate the validity of the modeled dual-systems factors, *a priori* hypotheses regarding the neural underpinnings of top-down and bottom-up constructs were tested by estimating genetic correlations between the dual-systems factors and relevant neuroimaging phenotypes. For instance, as hypothesized based on prior research and theory (Dalley & Robbins, 2017; Pan et al., 2021; Shulman et al., 2016), TH, especially in frontal regions, was negatively correlated in the present study with (lack of) self-control reflecting overlap in genetic variation associated with thinner frontocortical regions and diminished self-control (max |*r*_*g*_|=.43). This mirrors results from previous neuroimaging studies suggesting negative associations between frontocortical thickness and lack of self-control IPTs (Holmes, Hollinshead, Roffman, Smoller, & Buckner, 2016; Kaag et al., 2014; Kubera et al., 2018; Schilling et al., 2012). Additionally, the current findings compliment prior research with these TH GWAS data reporting a positive correlation between TH in the precentral gyrus and general cognitive functioning (Grasby et al., 2020). Together, these lines of evidence imply that associations between reduced frontocortical functioning and diminished self-control may be partially explained by a common underlying genetic basis, thus lending further support to the interpretation of the modeled latent (lack of) self-control factor as a top-down construct.

However, some findings ran contrary to the hypothesized neurobiology of the dual-systems model. The observed positive genetic correlations between sensation seeking and SA across a number of cortical regions were unexpected as dual-systems theory contends that sensation seeking, as a bottom-up construct, is primarily localized to subcortical structures (Steinberg et al., 2008). Similarly, previous research would partially situate urgency in the morphology of subcortical structures (e.g., amygdala, basal ganglia; Chester et al., 2016; Cyders et al., 2015; Halcomb, Argyriou, & Cyders, 2019). Nonetheless, urgency was significantly associated with a single neuroimaging phenotype following FDR correction: rostral middle frontal TH (*r*_*g*_=-.39), which replicates prior phenotypic studies of neuroimaging correlates of urgency (Cyders, Dzemidzic, et al., 2014; Cyders et al., 2015; Muhlert & Lawrence, 2015). In the current study, all dual-systems constructs were uncorrelated with subcortical structural volume. While prior evidence of associations between bottom-up constructs and subcortical structural volume has been limited to mixed findings of generally small effect (e.g., Holmes et al., 2016; Owens et al., 2020, 2022), functional neuroimaging studies have demonstrated a wider range of findings including unique patterns of connectivity across frontostriatal pathways (Burnette et al., 2019; Demidenko, Huntley, Weigard, Keating, & Beltz, 2022; Hawes et al., 2017; Um, Hummer, & Cyders, 2020; Zhu, Cortes, Mathur, Tomasi, & Momenan, 2017). In aggregate, current study findings suggest further research validating bottom-up constructs from neuroimaging and genetic perspectives is needed, though the consistency reported here with prior research and ongoing efforts to increase neuroimaging sample sizes highlight the promise of this approach (Marek et al., 2022).

In general, the mixed evidence reported here likely reflects a blend of (A) the complex genetic architecture of these traits and their neurogenetic associations with underlying neurobiological structure and function, and (B) potentially limited ability to separate or distill these factors into precise top-down and bottom-up features. Criticisms of the dual-systems model as overly simplified have noted that, while dual-systems constructs are theoretically and empirically separable (Duckworth & Steinberg, 2015; Shulman, Harden et al., 2016), the underlying neurobiology is likely dynamic and multifaceted (Casey, Galván, & Somerville, 2016; Pfiefer & Allen, 2012). Moreover, given that these constructs tend to be highly correlated (Steinberg et al., 2008), as was the case in the current study, investigations of genetic correlations between one of these constructs (e.g., sensation seeking) and any neurobiological analogue in isolation (e.g., neuroimaging phenotype) will be contaminated by the contribution of the unmeasured construct (e.g., [lack of] self-control; Shulman et al., 2016). Nonetheless, the current study represents a first effort to examine genetic interplay between these phenotypes in depth, and therefore, may serve as a benchmark for future studies assessing evidence for these neurogenetic associations.

### Limitations and Future Directions

The current study is not without limitations. First, limited sample sizes likely hindered analyses of the (lack of) self-control and urgency traits, demonstrating the need for larger IPT GWAS (e.g., Becker et al., 2021; Sanchez-Roige et al., 2022). Second, and equally important, is the exclusion of non-European ancestry samples from the present study. Crucially, non-European ancestry groups are extremely underrepresented in extant GWAS studies (Mills & Rahal, 2019, 2020), and addressing this limitation is of dire importance for leveling health disparities across groups (Peterson et al., 2019). Third, GWAS data were available only at the level of IPT subscales, rather than at the item-level, resulting in a small number of appropriate indicators for each genomic factor. Multivariate GWAS model construction using item-level summary statistics (e.g., Mallard, Savage, et al., 2022) would allow for the analysis of more complex models.

## Conclusion

Results of the current study suggest that a dual-systems model of the genetic architecture of IPTs is reasonably validated through genomic structural equation modeling, and factors derived from this model show disparate associations with relevant neuroimaging phenotypes. As such, this study serves as an important first step in defining the shared and unique genomic and neurobiological correlates of dual-systems constructs and underscores the importance of using imaging genetics to further elucidate neurobiological substrates underlying genetic overlap between complex traits (Bogdan et al., 2017).

## Data Availability

The full GWAS summary statistics for the 23andMe discovery data set will be made available through 23andMe to qualified researchers under an agreement with 23andMe that protects the privacy of the 23andMe participants. Please visit https://research.23andme.com/collaborate/#dataset-access/ for more information and to apply to access the data.

## Acknowledgements

This study made use of summary statistics data from a number of sources which we wish to acknowledge. First, we thank Susan Service, MSc, and Nelson Freimer, MD (University of California, Los Angeles), for providing TCI summary statistics from Service et al. (2012) for which compensation was not received. Second, this study made use of GWAS summary statistics data from 23andMe, Inc. (Sunnyvale, CA). We thank the 23andMe research participants and employees for making this work possible. Second, this research used summary data from UKB, a population-based sample of participants whose contributions we gratefully acknowledge. Finally, this study also made use of data generated by the UK10K Consortium, derived from samples from UK10K_COHORT_IMPUTATION REL-2012-06-02 (EGAD00001000776). A full list of the investigators who contributed to the generation of the data is available from www.UK10K.org. Funding for UK10K was provided by the Wellcome Trust under award WT091310.

All secondary data analysis of GWAS summary statistics and reference panels were considered exempt by the Institutional Review Board at the University of Missouri. The computations for all analyses were performed on the high-performance computing infrastructure provided by Research Computing Support Services and in part by the National Science Foundation under grant number CNS-1429294 at the University of Missouri, Columbia, MO. DOI: https://doi.org/10.32469/10355/69802

## References

Akaike, H. (1974). A new look at the statistical model identification. IEEE Transactions on Automatic Control, 19(6), 716–723. http://doi.org/10.1109/TAC.1974.1100705

Beauchaine, T. P., Zisner, A. R., & Sauder, C. L. (2017). Trait impulsivity and the externalizing spectrum. Annual Review of Clinical Psychology, 13, 343–368. https://doi.org/10.1146/annurev-clinpsy-021815-093253

Becker, J., Burik, C., Goldman, G., Wang, N., Jayashankar, H., Bennett, M., Belsky, D. W., Linnér, R. K., Ahlskog, R., Kleinman, A., Hinds, D. A., and Me Research Group, Caspi, A., Corcoran, D. L., Moffitt, T. E., Poulton, R., Sugden, K., Williams, B. S., Harris, K. M., Steptoe, A., … Okbay, A. (2021). Resource profile and user guide of the Polygenic Index Repository. Nature Human Behaviour, 5(12), 1744–1758. https://doi.org/10.1038/s41562-021-01119-3

Bentler, P. M. (1990). Comparative fit indexes in structural models. Psychological Bulletin, 107(2), 238–246. https://doi.org/10.1037/0033-2909.107.2.238

Bentler, P. M. (1995). EQS structural equations program manual. Encino, CA: Multivariate Software.

Bezdjian, S., Baker, L. A., & Tuvblad, C. (2011). Genetic and environmental influences on impulsivity: A meta-analysis of twin, family and adoption studies. Clinical Psychology Review, 31(7), 1209–1223. https://doi.org/10.1016/j.cpr.2011.07.005

Billieux, J., Heeren, A., Rochat, L., Maurage, P., Bayard, S., Bet, R., Besche-Richard, C., Challet-Bouju, G., Carré, A., Devos, G., Flayelle, M., Gierski, F., Grall-Bronnec, M., Kern, L., Khazaal, Y., Lançon, C., Lannoy, S., Michael, G. A., Raffard, S., Romo, L., … Baggio, S. (2021). Positive and negative urgency as a single coherent construct: Evidence from a large-scale network analysis in clinical and non-clinical samples. Journal of Personality, 89(6), 1252–1262. https://doi.org/10.1111/jopy.12655

Bogdan, R., Salmeron, B. J., Carey, C. E., Agrawal, A., Calhoun, V. D., Garavan, H., Hariri, A. R., Heinz, A., Hill, M. N., Holmes, A., Kalin, N. H., & Goldman, D. (2017). Imaging genetics and genomics in psychiatry: a critical review of progress and potential. Biological Psychiatry, 82(3), 165–175. https://doi.org/10.1016/j.biopsych.2016.12.030

Bulik-Sullivan, B., Finucane, H. K., Anttila, V., Gusev, A., Day, F. R., Loh, P. R., Duncan, L., Perry, J. R. B., Patterson, N., Robinson, E. B., Daly, M. J., Price, A. L., & Neale, B. M. (2015). An atlas of genetic correlations across human diseases and traits. Nature Genetics, 47(11), 1236–1241. https://doi.org/10.1038/ng.3406

Burnette, E. M., Grodin, E. N., Lim, A. C., MacKillop, J., Karno, M. P., & Ray, L. A. (2019). Association between impulsivity and neural activation to alcohol cues in heavy drinkers. Psychiatry Research Neuroimaging, 293, 110986. https://doi.org/10.1016/j.pscychresns.2019.110986

Carver, C. S., & Johnson, S. L. (2018). Impulsive reactivity to emotion and vulnerability to psychopathology. The American Psychologist, 73(9), 1067–1078. https://doi.org/10.1037/amp0000387

Casey, B. J., Galván, A., & Somerville, L. H. (2016). Beyond simple models of adolescence to an integrated circuit-based account: A commentary. Developmental Cognitive Neuroscience, 17, 128–130. https://doi.org/10.1016/j.dcn.2015.12.006

Casey, B. J., Getz, S., & Galvan, A. (2008). The adolescent brain. Developmental Review, 28(1), 62–77. https://doi.org/10.1016/j.dr.2007.08.003

Chester, D. S., Lynam, D. R., Milich, R., Powell, D. K., Andersen, A. H., & DeWall, C. N. (2016). How do negative emotions impair self-control? A neural model of negative urgency. NeuroImage, 132, 43–50. https://doi.org/10.1016/j.neuroimage.2016.02.024

Cloninger C. R., Przybeck T. R., Svrakic D. M., Wetzel R. D. (1994) The Temperament and Character Inventory (TCI): A guide to its development and use. St. Louis, MO: Center for Psychobiology of Personality.

Cloninger, C. R. (1987). A systematic method for clinical description and classification of personality variants. A proposal. Archives of General Psychiatry, 44(6), 573–588. https://doi.org/10.1001/archpsyc.1987.01800180093014

Congdon, E., & Canli, T. (2005). The endophenotype of impulsivity: Reaching consilience through behavioral, genetic, and neuroimaging approaches. Behavioral and Cognitive Neuroscience Reviews, 4(4), 262–281. https://doi.org/10.1177/1534582305285980

Creswell, K. G., Wright, A. G. C., Flory, J. D., Skrzynski, C. J., & Manuck, S. B. (2019). Multidimensional assessment of impulsivity-related measures in relation to externalizing behaviors. Psychological Medicine, 49(10), 1678–1690. https://doi.org/10.1017/S0033291718002295

Cyders, M. A., Coskunpinar, A., & VanderVeen, J. D. (2016). Urgency: A common transdiagnostic endophenotype for maladaptive risk taking. In V. Zeigler-Hill & D. K. Marcus (Eds.), The dark side of personality: Science and practice in social, personality, and clinical psychology (pp. 157–188). American Psychological Association. https://doi.org/10.1037/14854-009

Cyders, M. A., Dzemidzic, M., Eiler, W. J., Coskunpinar, A., Karyadi, K., & Kareken, D. A. (2015). Negative urgency and ventromedial prefrontal cortex responses to alcohol cues: fMRI evidence of emotion-based impulsivity. Alcoholism, Clinical and Experimental Research, 38(2), 409–417. https://doi.org/10.1111/acer.12266

Cyders, M. A., Dzemidzic, M., Eiler, W. J., Coskunpinar, A., Karyadi, K. A., & Kareken, D. A. (2015). Negative urgency mediates the relationship between amygdala and orbitofrontal cortex activation to negative emotional stimuli and general risk-taking. Cerebral Cortex, 25(11), 4094–4102. https://doi.org/10.1093/cercor/bhu123

Cyders, M. A., Littlefield, A. K., Coffey, S., & Karyadi, K. A. (2014). Examination of a short English version of the UPPS-P Impulsive Behavior Scale. Addictive Behaviors, 39(9), 1372–1376. https://doi.org/10.1016/j.addbeh.2014.02.013

Dalley, J. W., & Robbins, T. W. (2017). Fractionating impulsivity: Reuropsychiatric implications. Nature Reviews Neuroscience, 18(3), 158–171. https://doi.org/10.1038/nrn.2017.8

Demidenko, M. I., Huntley, E. D., Weigard, A. S., Keating, D. P., & Beltz, A. M. (2022). Neural heterogeneity underlying late adolescent motivational processing is linked to individual differences in behavioral sensation seeking. Journal of Neuroscience Research, 100(3), 762–779. https://doi.org/10.1002/jnr.25005

Desikan, R. S., Ségonne, F., Fischl, B., Quinn, B. T., Dickerson, B. C., Blacker, D., Buckner, R. L., Dale, A. M., Maguire, R. P., Hyman, B. T., Albert, M. S., & Killiany, R. J. (2006). An automated labeling system for subdividing the human cerebral cortex on MRI scans into gyral based regions of interest. NeuroImage, 31(3), 968–980. https://doi.org/10.1016/j.neuroimage.2006.01.021

Duckworth, A., & Steinberg, L. (2015). Unpacking self-control. Child Development Perspectives, 9(1), 32–37. https://doi.org/10.1111/cdep.12107

Duell, N., Steinberg, L., Chein, J., Al-Hassan, S. M., Bacchini, D., Lei, C., Chaudhary, N., Di Giunta, L., Dodge, K. A., Fanti, K. A., Lansford, J. E., Malone, P. S., Oburu, P., Pastorelli, C., Skinner, A. T., Sorbring, E., Tapanya, S., Uribe Tirado, L. M., & Alampay, L. P. (2016). Interaction of reward seeking and self-regulation in the prediction of risk taking: A cross-national test of the dual systems model. Developmental Psychology, 52(10), 1593–1605. https://doi.org/10.1037/dev0000152

Ellingson, J. M., Slutske, W. S., Vergés, A., Littlefield, A. K., Statham, D. J., & Martin, N. G. (2018). A multivariate behavior genetic investigation of dual-systems models of alcohol involvement. Journal of Studies on Alcohol and Drugs, 79(4), 617–626. https://doi.org/10.15288/JSAD.2018.79.617

Ellingson, J. M., Vergés, A., Littlefield, A. K., Martin, N. G., & Slutske, W. S. (2013). Are bottom-up and top-down traits in dual-systems models of risky behavior genetically distinct? Behavior Genetics, 43(6), 480–490. https://doi.org/10.1007/s10519-013-9615-9

Ernst, M. (2014). The triadic model perspective for the study of adolescent motivated behavior. Brain and Cognition, 89, 104–111. https://doi.org/10.1016/j.bandc.2014.01.006

Evren, C., Durkaya, M., Evren, B., Dalbudak, E., & Cetin, R. (2012). Relationship of relapse with impulsivity, novelty seeking and craving in male alcohol-dependent inpatients. Drug and Alcohol Review, 31(1), 81–90. https://doi.org/10.1111/j.1465-3362.2011.00303.x

Fischer, S., Smith, G. T., & Cyders, M. A. (2008). Another look at impulsivity: A meta-analytic review comparing specific dispositions to rash action in their relationship to bulimic symptoms. Clinical Psychology Review, 28(8), 1413–1425. https://doi.org/10.1016/j.cpr.2008.09.001

Friedman, N. P., Hatoum, A. S., Gustavson, D. E., Corley, R. P., Hewitt, J. K., & Young, S. E. (2020). Executive functions and impulsivity are genetically distinct and independently predict psychopathology: Results from two adult twin studies. Clinical Psychological Science, 8(3), 519–538. https://doi.org/10.1177/2167702619898814

Grasby, K. L., Jahanshad, N., Painter, J. N., Colodro-Conde, L., Bralten, J., Hibar, D. P., Lind, P. A., Pizzagalli, F., Ching, C., McMahon, M., Shatokhina, N., Zsembik, L., Thomopoulos, S. I., Zhu, A. H., Strike, L. T., Agartz, I., Alhusaini, S., Almeida, M., Alnæs, D., Amlien, I. K., …Enhancing NeuroImaging Genetics through Meta-Analysis Consortium (ENIGMA)—Genetics working group (2020). The genetic architecture of the human cerebral cortex. Science, 367(6484), eaay6690. https://doi.org/10.1126/science.aay6690

Grotzinger, A. D., Rhemtulla, M., de Vlaming, R., Ritchie, S. J., Mallard, T. T., Hill, W. D., Ip, H. F., Marioni, R. E., McIntosh, A. M., Deary, I. J., Koellinger, P. D., Harden, K. P., Nivard, M. G., & Tucker-Drob, E. M. (2019). Genomic structural equation modelling provides insights into the multivariate genetic architecture of complex traits. Nature Human Behaviour, 3(5), 513–525. https://doi.org/10.1038/s41562-019-0566-x

Gustavson, D. E., Franz, C. E., Kremen, W. S., Carver, C. S., Corley, R. P., Hewitt, J. K., & Friedman, N. P. (2019). Common genetic influences on impulsivity facets are related to goal management, psychopathology, and personality. Journal of Research in Personality, 79, 161–175. https://doi.org/10.1016/j.jrp.2019.03.009

Gustavson, D. E., Friedman, N. P., Fontanillas, P., Elson, S. L., 23andMe Research Team, Palmer, A. A., & Sanchez-Roige, S. (2020). The latent genetic structure of impulsivity and its relation to internalizing psychopathology. Psychological Science, 31(8), 1025– 1035. https://doi.org/10.1177/0956797620938160

Halcomb, M., Argyriou, E., & Cyders, M. A. (2019). Integrating preclinical and clinical models of negative urgency. Frontiers in Psychiatry, 10, 324. https://doi.org/10.3389/fpsyt.2019.00324

Hall, M. H., & Smoller, J. W. (2010). A new role for endophenotypes in the GWAS era: Functional characterization of risk variants. Harvard Review of Psychiatry, 18(1), 67–74. https://doi.org/10.3109/10673220903523532

Harden, K. P., Kretsch, N., Mann, F. D., Herzhoff, K., Tackett, J. L., Steinberg, L., & Tucker-Drob, E. M. (2017). Beyond dual systems: A genetically-informed, latent factor model of behavioral and self-report measures related to adolescent risk-taking. Developmental Cognitive Neuroscience, 25, 221–234. https://doi.org/10.1016/j.dcn.2016.12.007

Hawes, S. W., Chahal, R., Hallquist, M. N., Paulsen, D. J., Geier, C. F., & Luna, B. (2017). Modulation of reward-related neural activation on sensation seeking across development. NeuroImage, 147, 763–771. https://doi.org/10.1016/j.neuroimage.2016.12.020

Holmes, A. J., Hollinshead, M. O., Roffman, J. L., Smoller, J. W., & Buckner, R. L. (2016). Individual differences in cognitive control circuit anatomy link sensation seeking, impulsivity, and substance use. The Journal of Neuroscience, 36(14), 4038–4049. https://doi.org/10.1523/JNEUROSCI.3206-15.2016

Hur, Y. M., & Bouchard, T. J. (1997). The genetic correlation between impulsivity and sensation seeking traits. Behavior Genetics, 27(5), 455–463. https://doi.org/10.1023/A:1025674417078

Jiménez-Murcia, S., Granero, R., Giménez, M., Del Pino-Gutiérrez, A., Mestre-Bach, G., Mena-Moreno, T., Moragas, L., Baño, M., Sánchez-González, J., de Gracia, M., Baenas-Soto, I., Contaldo, S. F., Valenciano-Mendoza, E., Mora-Maltas, B., López-González, H., Menchón, J. M., & Fernández-Aranda, F. (2020). Contribution of sex on the underlying mechanism of the gambling disorder severity. Scientific Reports, 10(1), 18722. https://doi.org/10.1038/s41598-020-73806-6

Johnson, S. L., Carver, C. S., & Joormann, J. (2013). Impulsive responses to emotion as a transdiagnostic vulnerability to internalizing and externalizing symptoms. Journal of Affective Disorders, 150(3), 872–878. https://doi.org/10.1016/j.jad.2013.05.004

Johnson, S. L., Elliott, M. V., & Carver, C. S. (2020). Impulsive responses to positive and negative emotions: Parallel neurocognitive correlates and their implications. Biological Psychiatry, 87(4), 338–349. https://doi.org/10.1016/j.biopsych.2019.08.018

Jonas, K. G., & Markon, K. E. (2014). A meta-analytic evaluation of the endophenotype hypothesis: effects of measurement paradigm in the psychiatric genetics of impulsivity. Journal of Abnormal Psychology, 123(3), 660–675. https://doi.org/10.1037/a0037094

Kaag, A. M., Crunelle, C. L., van Wingen, G., Homberg, J., van den Brink, W., & Reneman, L. (2014). Relationship between trait impulsivity and cortical volume, thickness and surface area in male cocaine users and non-drug using controls. Drug and Alcohol Dependence, 144, 210–217. https://doi.org/10.1016/j.drugalcdep.2014.09.016

Kendler, K. S., & Neale, M. C. (2010). Endophenotype: A conceptual analysis. Molecular Psychiatry, 15(8), 789–797. https://doi.org/10.1038/mp.2010.8

Klein, A., & Tourville, J. (2012). 101 labeled brain images and a consistent human cortical labeling protocol. Frontiers in Neuroscience, 6, 171. https://doi.org/10.3389/fnins.2012.00171

Kubera, K. M., Schmitgen, M. M., Maier-Hein, K. H., Thomann, P. A., Hirjak, D., & Wolf, R. C. (2018). Differential contributions of cortical thickness and surface area to trait impulsivity in healthy young adults. Behavioural Brain Research, 350, 65–71. https://doi.org/10.1016/j.bbr.2018.05.006

Linnér, R. K., Biroli, P., Kong, E., Meddens, S., Wedow, R., Fontana, M. A., Lebreton, M., Tino, S. P., Abdellaoui, A., Hammerschlag, A. R., Nivard, M. G., Okbay, A., Rietveld, C. A., Timshel, P. N., Trzaskowski, M., Vlaming, R., Zünd, C. L., Bao, Y., Buzdugan, L., Caplin, A. H., … Beauchamp, J. P. (2019). Genome-wide association analyses of risk tolerance and risky behaviors in over 1 million individuals identify hundreds of loci and shared genetic influences. Nature Genetics, 51(2), 245–257. https://doi.org/10.1038/s41588-018-0309-3

Lopez-Vergara, H. I., Spillane, N. S., Merrill, J. E., & Jackson, K. M. (2016). Developmental trends in alcohol use initiation and escalation from early to middle adolescence: Prediction by urgency and trait affect. Psychology of Addictive, 30(5), 578–587. https://doi.org/10.1037/adb0000173

Mallard, T. T., Linnér, R. K., Grotzinger, A. D., Sanchez-Roige, S., Seidlitz, J., Okbay, A., de Vlaming, R., Meddens, S. F. W., Palmer, A. A., Davis, L. K., Tucker-Drob, E. M., Kendler, K. S., Keller, M. C., Koellinger, P. D. & Harden, K. P. (2022). Multivariate GWAS of psychiatric disorders and their cardinal symptoms reveal two dimensions of cross-cutting genetic liabilities. Cell Genomics, 2(6), 100140. https://doi.org/10.1016/j.xgen.2022.100140

Mallard, T. T., Savage, J. E., Johnson, E. C., Huang, Y., Edwards, A. C., Hottenga, J. J., Grotzinger, A. D., Gustavson, D. E., Jennings, M. V., Anokhin, A., Dick, D. M., Edenberg, H. J., Kramer, J. R., Lai, D., Meyers, J. L., Pandey, A. K., Harden, K. P., Nivard, M. G., de Geus, E., Boomsma, D. I., … Sanchez-Roige, S. (2022). Item-level genome-wide association study of the Alcohol Use Disorders Identification Test in three population-based cohorts. The American Journal of Psychiatry, 179(1), 58–70. https://doi.org/10.1176/appi.ajp.2020.20091390

Marek, S., Tervo-Clemmens, B., Calabro, F. J., Montez, D. F., Kay, B. P., Hatoum, A. S., Donohue, M. R., Foran, W., Miller, R. L., Hendrickson, T. J., Malone, S. M., Kandala, S., Feczko, E., Miranda-Dominguez, O., Graham, A. M., Earl, E. A., Perrone, A. J., Cordova, M., Doyle, O., Moore, L. A., … Dosenbach, N. (2022). Reproducible brain-wide association studies require thousands of individuals. Nature, 603(7902), 654–660. https://doi.org/10.1038/s41586-022-04492-9

Matsuo, K., Nicoletti, M., Nemoto, K., Hatch, J. P., Peluso, M. A., Nery, F. G., & Soares, J. C. (2009). A voxel-based morphometry study of frontal gray matter correlates of impulsivity. Human Brain Mapping, 30(4), 1188–1195. https://doi.org/10.1002/hbm.20588

Miller, A. P., & Gizer, I. R. (2023). Neurogenetic and multi-omic sources of overlap among sensation seeking, alcohol consumption, and alcohol use disorder. Manuscript in preparation.

Mills, M. C., & Rahal, C. (2019). A scientometric review of genome-wide association studies. Communications Biology, 2, 9. https://doi.org/10.1038/s42003-018-0261-x

Morean, M. E., DeMartini, K. S., Leeman, R. F., Pearlson, G. D., Anticevic, A., Krishnan-Sarin, S., Krystal, J. H., & O’Malley, S. S. (2014). Psychometrically improved, abbreviated versions of three classic measures of impulsivity and self-control. Psychological Assessment, 26(3), 1003–1020. https://doi.org/10.1037/pas0000003

Mowinckel, A. M., & Vidal-Piñeiro, D. (2020). Visualization of brain statistics with R packages ggseg and ggseg3d. Advances in Methods and Practices in Psychological Science, 3(4), 466–483. https://doi.org/10.1177/2515245920928009

Muhlert, N., & Lawrence, A. D. (2015). Brain structure correlates of emotion-based rash impulsivity. NeuroImage, 115, 138–146. https://doi.org/10.1016/j.neuroimage.2015.04.061

Owens, M. M., Hyatt, C. S., Gray, J. C., Miller, J. D., Lynam, D. R., Hahn, S., Allgaier, N., Potter, A., & Garavan, H. (2020). Neuroanatomical correlates of impulsive traits in children aged 9 to 10. Journal of Abnormal Psychology, 129(8), 831–844. https://doi.org/10.1037/abn0000627

Owens, M. M., Hyatt, C. S., Xu, H., Thompson, M. F., Miller, J. D., Lynam, D. R., MacKillop, J., & Gray, J. C. (2022). Replicability of the neuroanatomical correlates of impulsive personality traits in the ABCD study. PsyArXiv. https://doi.org/10.31234/osf.io/u3bxc

Pan, N., Wang, S., Zhao, Y., Lai, H., Qin, K., Li, J., Biswal, B. B., Sweeney, J. A., & Gong, Q. (2021). Brain gray matter structures associated with trait impulsivity: A systematic review and voxel-based meta-analysis. Human Brain Mapping, 42(7),2214– 2235. https://doi.org/10.1002/hbm.25361

Patton, J., Stanford, M., & S. Barratt E. (1995). Factor structure of the Barratt Impulsiveness Scale. Journal of Clinical Psychology, 51, 768–774. https://doi.org/10.1002/1097-4679(199511)51:63.0.CO;2-1

Peterson, R. E., Kuchenbaecker, K., Walters, R. K., Chen, C. Y., Popejoy, A. B., Periyasamy, S., Lam, M., Iyegbe, C., Strawbridge, R. J., Brick, L., Carey, C. E., Martin, A. R., Meyers, J. L., Su, J., Chen, J., Edwards, A. C., Kalungi, A., Koen, N., Majara, L., Schwarz, E., … Duncan, L. E. (2019). Genome-wide association studies in ancestrally diverse populations: Opportunities, methods, pitfalls, and recommendations. Cell, 179(3), 589– 603. https://doi.org/10.1016/j.cell.2019.08.051

Pfeifer, J. H., & Allen, N. B. (2012). Arrested development? Reconsidering dual-systems models of brain function in adolescence and disorders. Trends in Cognitive Sciences, 16(6), 322– 329. https://doi.org/10.1016/j.tics.2012.04.011

Reise, S. P., Moore, T. M., Sabb, F. W., Brown, A. K., & London, E. D. (2013). The Barratt Impulsiveness Scale-11: Reassessment of its structure in a community sample. Psychological Assessment, 25(2), 631–642. https://doi.org/10.1037/a0032161

Rüeger, S., McDaid, A., & Kutalik, Z. (2018). Evaluation and application of summary statistic imputation to discover new height-associated loci. PLoS Genetics, 14(5), e1007371. https://doi.org/10.1371/journal.pgen.1007371

Sanchez-Roige, S., Fontanillas, P., Elson, S. L., Gray, J. C., De Wit, H., MacKillop, J., & Palmer, A. A. (2019). Genome-wide association studies of impulsive personality traits (BIS-11 and UPPS-P) and drug experimentation in up to 22,861 adult research participants identify loci in the CACNA1I and CADM2 genes. Journal of Neuroscience, 39(13), 2562–2572. https://doi.org/10.1523/JNEUROSCI.2662-18.2019

Sanchez-Roige, S., Jennings, M. V., Thorpe, H. H., Mallari, J. E., van der Werf, L. C., Bianchi, S. B., Lee, C., Mallard, T. T., Barnes, S. A., Wu, J. Y., Barkley-Levenson, A. M., Boussaty, E. C., Snethlage, C. E., Schafer, D., Babic, Z., Winters, B. D., Watters, K. E., Biederer, T., Mackillop, J., … Palmer, A. A. (2022). CADM2 is implicated in impulsive personality and numerous other traits by genome-and phenome-wide association studies in humans, with further support from studies of CADM2 mutant mice. medRxiv. https://doi.org/10.1101/2022.01.29.22270095

Satizabal, C. L., Adams, H., Hibar, D. P., White, C. C., Knol, M. J., Stein, J. L., Scholz, M., Sargurupremraj, M., Jahanshad, N., Roshchupkin, G. V., Smith, A. V., Bis, J. C., Jian, X., Luciano, M., Hofer, E., Teumer, A., van der Lee, S. J., Yang, J., Yanek, L. R., Lee, T. V., … Ikram, M. A. (2019). Genetic architecture of subcortical brain structures in 38,851 individuals. Nature Genetics, 51(11), 1624–1636. https://doi.org/10.1038/s41588-019-0511-y

Savvidou, L. G., Fagundo, A. B., Fernández-Aranda, F., Granero, R., Claes, L., Mallorquí-Baqué, N., Verdejo-García, A., Steiger, H., Israel, M., Moragas, L., Del Pino-Gutiérrez, A., Aymamí, N., Gómez-Peña, M., Agüera, Z., Tolosa-Sola, I., La Verde, M., Aguglia, E., Menchón, J. M., & Jiménez-Murcia, S. (2017). Is gambling disorder associated with impulsivity traits measured by the UPPS-P and is this association moderated by sex and age? Comprehensive Psychiatry, 72, 106–113. https://doi.org/10.1016/j.comppsych.2016.10.005

Schilling, C., Kühn, S., Romanowski, A., Schubert, F., Kathmann, N., & Gallinat, J. (2012). Cortical thickness correlates with impulsiveness in healthy adults. NeuroImage, 59(1), 824–830. https://doi.org/10.1016/j.neuroimage.2011.07.058

Service, S. K., Verweij, K. J. H., Lahti, J., Congdon, E., Ekelund, J., Hintsanen, M., Räikkönen, K., Lehtimäki, T., Kähönen, M., Widen, E., Taanila, A., Veijola, J., Heath, A. C., Madden, P. A. F., Montgomery, G. W., Sabatti, C., Järvelin, M. R., Palotie, A., Raitakari, O., … Freimer, N. B. (2012). A genome-wide meta-analysis of association studies of Cloninger’s Temperament Scales. Translational Psychiatry, 2, 1–9. https://doi.org/10.1038/tp.2012.37

Shulman, E. P., Harden, K. P., Chein, J. M., & Steinberg, L. (2016). The development of impulse control and sensation-seeking in adolescence: Independent or interdependent processes? Journal of Research on Adolescence, 26(1), 37–44. https://doi.org/10.1111/jora.12181

Shulman, E. P., Smith, A. R., Silva, K., Icenogle, G., Duell, N., Chein, J., & Steinberg, L. (2016). The dual systems model: Review, reappraisal, and reaffirmation. Developmental Cognitive Neuroscience, 17, 103–117. https://doi.org/10.1016/j.dcn.2015.12.010

Smith, S. M., Douaud, G., Chen, W., Hanayik, T., Alfaro-Almagro, F., Sharp, K., & Elliott, L. T. (2021). An expanded set of genome-wide association studies of brain imaging phenotypes in UK Biobank. Nature Neuroscience, 24(5), 737–745. https://doi.org/10.1038/s41593-021-00826-4

Stautz, K., & Cooper, A. (2013). Impulsivity-related personality traits and adolescent alcohol use: A meta-analytic review. Clinical Psychology Review, 33(4), 574–592.

Steinberg, L., Albert, D., Cauffman, E., Banich, M., Graham, S., & Woolard, J. (2008). Age differences in sensation seeking and impulsivity as indexed by behavior and self-report: Evidence for a dual systems model. Developmental Psychology, 44(6), 1764.

Strickland, J. C., & Johnson, M. W. (2021). Rejecting impulsivity as a psychological construct: A theoretical, empirical, and sociocultural argument. Psychological Review, 128(2), 336.

The 1000 Genomes Project Consortium. (2015). A global reference for human genetic variation. Nature, 526(7571), 68–74. https://doi.org/10.1038/nature15393

Um, M., Hummer, T. A., & Cyders, M. A. (2020). Relationship of negative urgency to cingulo-insular and cortico-striatal resting state functional connectivity in tobacco use. Brain Imaging and Behavior, 14(5), 1921–1932. https://doi.org/10.1007/s11682-019-00136-1

Verdejo-Garcia, A., & Albein-Urios, N. (2021). Impulsivity traits and neurocognitive mechanisms conferring vulnerability to substance use disorders. Neuropharmacology, 183, 108402. https://doi.org/10.1016/j.neuropharm.2020.108402

Vonmoos, M., Hulka, L. M., Preller, K. H., Jenni, D., Schulz, C., Baumgartner, M. R., & Quednow, B. B. (2013). Differences in self-reported and behavioral measures of impulsivity in recreational and dependent cocaine users. Drug and Alcohol Dependence, 133(1), 61–70. https://doi.org/10.1016/j.drugalcdep.2013.05.032

Whiteside, S. P., & Lynam, D. R. (2001). The Five Factor Model and impulsivity: Using a structural model of personality to understand impulsivity. Personality and Individual Differences, 30(4), 669–689. https://doi.org/10.1016/S0191-8869(00)00064-7

Zhu, X., Cortes, C. R., Mathur, K., Tomasi, D., & Momenan, R. (2017). Model-free functional connectivity and impulsivity correlates of alcohol dependence: A resting-state study. Addiction Biology, 22(1), 206–217. https://doi.org/10.1111/adb.12272

