## Supplementary Methods for "Dual-systems models of the genetic architecture of impulsive personality traits: Neurogenetic evidence of distinct but related factors"

#### *Impulsive Personality Trait GWAS*

**Genotyping, Imputation, and Quality Control**. Genome-wide association study (GWAS) summary statistics for impulsive personality trait (IPT) phenotypes were obtained from three primary sources: the UK Biobank (UKB; Bycroft et al., 2018), direct-to-consumer genetics company 23andMe, Inc. (Sunnyvale, CA), and a meta-analytic sample comprised of the Northern Finland Birth Cohort, the Cardiovascular Risk in Young Finns Study, the Helsinki Birth Cohort Study, and the Australian twin registry which were provided upon request for additional IPT phenotypes (Service et al., 2012).

Briefly, 23andMe samples were genotyped using Illumina (San Diego, CA) platforms (Illumina HumanHap550+ Bead chip V1 V2, OmniExpress+ Bead chip V3, Custom array V4). Following quality control (QC) procedures (e.g., call rate > 95%, Hardy–Weinberg equilibrium [HWE] $P < 1 \times 10^{-20}$, restriction to unrelated individuals using identity-by-descent [IBD] approaches), genotypes were imputed against Phase 1 v3 of 1000 Genomes Project haplotypes (The 1000 Genomes Project Consortium, 2010) using Minimac2 (Fuchsberger, Abecasis, & Hinds, 2015). Imputation resulted in > 9 million high-quality SNPs ($r^2 \geq .5$) for association analyses of each trait. QC of genetic variants, imputation, and genome-wide analyses were performed by 23andMe, and summary statistics of IPT phenotypes were provided by request (see Sanchez-Roige et al., 2019 for a full account of these methods).

UKB samples were genotyped using one of two platforms: the Applied Biosystems UK BiLEVE Axiom and Applied Biosystems UK Biobank Axiom Arrays by Affymetrix (Santa Clara, CA). Following QC, genotypes were imputed to combined Haplotype Reference Consortium (HRCr1.1; McCarthy et al., 2016) and merged UK10K and 1000 Genomes Phase 3 reference panels (Huang et al., 2015) using IMPUTE4 (Howie, Fuchsberger, Stephens, Marchini, & Abecasis, 2012). For greater detail on genotyping, QC, and imputation of all UKB data and smaller replication samples for the 'risk taking' phenotype, see Bycroft et al. (2018) and Linnér et al. (2019), respectively. UKB and replication sample GWAS summary statistics for the 'risk taking' phenotype were obtained from the Social Science Genetic Association Consortium (SSGAC; https://thessgac.com/).

GWAS data from the Service et al. meta-analytic samples were genotyped across various Illumina and Affymetrix platforms. QC procedures were conducted for variants and samples based on missing data, call rates, sex discrepancy, ancestral outliers, and HWE ($P < 1 \times 10^{-5}$). QCed genotype data were imputed to the European panel of the HapMap2 reference sample (The International HapMap Consortium, 2007), separately by cohort, using the Markov Chain Haplotyper (MaCH; Li et al., 2009) prior to sample-size weighted meta-analysis in METAL (Willer, Li, & Abecasis, 2010). Following imputation and QC procedures, 1,252,387 high-quality ($r^2 > .3$) SNPs across cohorts were available for association analyses. Given the comparatively low number of tested variants in this study (i.e., versus > 5 million in more recent studies), summary statistics imputation was conducted for the Service et al. GWAS of TCI subscales as described in the *Summary Statistics Imputation* section below.

**Primary IPT Measures.** GWAS IPT phenotypes were primarily measured using the 20-item brief version of the UPPS-P Impulsive Behavior Scale (Cyders, Littlefield, Coffey, & Karyadi, 2014), the Barratt Impulsiveness Scale (BIS-11; Patton, Stanford, & Barratt, 1995), and Cloninger's Temperament and Character Inventory (TCI; Cloninger, Przybeck, Svrakic, & Wetzel, 1994). The 20-item brief version of the UPPS-P measures dimensions of impulsive personality using five subscales: *(lack of) premeditation* ("I usually think carefully before doing anything"), *(lack of) perseverance* ("I finish what I start"), *negative urgency* ("When I am upset I often act without thinking"), *positive urgency* ("I tend to lose control when I am in a great mood"), and *sensation seeking* ("I would enjoy the sensation of skiing very fast down a high mountain slope"). Each subscale is assessed by four 4-point Likert scale items (1 = "Agree strongly", 2 = "Agree somewhat", 3 = "Disagree somewhat", 4 = "Disagree strongly"). Some items are reverse coded, and subscales are scored such that greater scores represent greater levels of impulsivity for each trait. Internal consistencies were acceptable for each subscale phenotype assessed in the 23andMe GWAS sample (Cronbach's $\alpha \geq .70$; Sanchez-Roige et al., 2019).

The BIS-11 is a 30-item measure designed to assess three domains of impulsive personality: *attentional impulsivity* ("I don't pay attention"), *motor impulsivity* ("I do things without thinking"), and *non-planning impulsivity* ("I am self-controlled"). Each domain contains eight to 11 items measured on a 4-point Likert scale (1 = "Rarely/never", 2 = "Occasionally", 3 = "Often", 4 = "Almost always/always"). Some items are reverse coded, and items are scored such that greater scores represent greater levels of impulsivity for each domain. A total score is also often calculated by summing items across all subscales. Internal consistencies for domain scores assessed in the 23andMe GWAS sample (Sanchez-Roige et al., 2019) ranged from poor (motor impulsivity Cronbach's $\alpha = .54$) to acceptable (attentional impulsivity Cronbach's $\alpha = .74$; non-planning impulsivity Cronbach's $\alpha = .73$). Internal consistency of the total score was good (Cronbach's $\alpha = .83$).

The TCI is a 226-item measure containing binary (True/False) response options assessing four primary domains of temperament: *novelty seeking* ("I am much more reserved and controlled than most people"), *harm avoidance* ("I usually feel tense and worried when I have to do something new and unfamiliar"), *reward dependence* ("I like to please other people as much as I can"), and *persistence* ("I am usually so determined that I continued to work long after other people have given up"). Though each sample in the Service et al. TCI GWAS meta-analysis used slightly different item subsets and versions of the TCI, these four subscales were retained across each sample. Though internal consistencies of the temperament subscales for each meta-analytic sample were not reported by Service et al., in a previous report of internal consistencies of these subscales in the Northern Finland Birth Cohort values ranged from poor (persistence Cronbach's $\alpha = .55$) to good (harm avoidance Cronbach's $\alpha = .85$; Miettunen et al., 2004).

**Summary Statistics Imputation.** As noted above, GWAS data for TCI subscale phenotypes (novelty seeking and harm avoidance) were originally imputed to HapMap2 separately by cohort, using MaCH prior to sample-size weighted meta-analysis in METAL. Following QC procedures, 1,252,387 SNPs across cohorts were available for association analyses. To obtain similar sets of imputed variants across traits adequate for multivariate modeling of GWAS using GenomicSEM, summary statistics imputation was applied to impute tagged variants thus allowing for association tests with a range of variants comparable to more recent impulsivity GWAS (e.g., Sanchez-Roige et al., 2019). GWAS summary statistics for TCI novelty seeking and harm avoidance were imputed to a merged reference panel using the UK10K cohort (REL-2012-06-02; The UK10K Consortium, 2015) plus the European subsample of the 1000 Genomes Project Phase3 data (release 20131101.v5; The 1000 Genomes Project Consortium, 2015). This merged reference panel was accessed through the European Genome-Phenome Archive (EGAD00001000776). The combined sample size of this reference panel is 4,284.

To impute summary statistics for missing variants, the summary statistics imputation method implemented in the ssimp v0.5.6 software was used for SNPs with MAF ≥ 0.01 (Rüeger, McDaid, & Kutalik, 2018). Briefly, summary statistics imputation harnesses the linkage disequilibrium (LD) information estimated from the matched sequencing reference panels to directly impute meta-analysis summary statistics. To account for varying sample sizes across tag SNPs, the overlap between samples was modeled using dependent missingness (--missingness *dep*) which results in a lower RMSE of imputed statistics when sample size variance is low as is the case with the TCI summary statistics (Rüeger, McDaid, & Kutalik, 2018). SNPs with imputation quality scores of ($r^2$) < 0.7 were excluded following imputation, and the resulting summary statistics for TCI novelty seeking and harm avoidance contained test statistics for 6,039,602 and 6,041,944 SNPs respectively.

**Risk Taking GWAS Meta-Analysis**. A GWAS meta-analysis was conducted on risk tolerance (risk taking) phenotypes assessed via Likert-scale and binary items ("Would you describe yourself as someone who takes risks?") utilizing GWAS summary statistics for this phenotype from 23andMe ($N$ = 24,302) and the UKB cohort along with ten smaller replication samples (Linnér et al., 2019; $N$ = 466,571). For this risk taking GWAS meta-analysis, METAL was used to conduct a one-stage sample-size-weighted meta-analysis of the two non-overlapping cohort-level GWAS summary statistics for SNPs with MAF ≥ 0.01. Preliminary genetic correlation analyses confirmed a high degree of concordance between the 23andMe and Linnér et al. risk taking summary statistics: $r_g = .83$, $SE = .08$, $P = 1.78 \times 10^{-23}$. The resulting risk taking meta-analytic summary statistics ($N$ = 490,873) were used as an indicator GWAS for downstream GenomicSEM analyses (see Supplementary Figure 1 for quantile-quantile [Q-Q] plots of these meta-analytic results). Genomic control was not applied to METAL results for any trait as meta-analysis–level genomic control correction can be overly conservative for traits used in downstream LDSC-based analyses, generally yielding an LDSC intercept less than 1 (Bulik-Sullivan, Loh, et al., 2015; Lee, McGue, Iacono, & Chow, 2018).

### *Neuroimaging GWAS*

**Genotyping, Imputation, and Quality Control**. GWAS summary statistics for neuroimaging phenotypes were obtained from three primary sources: UKB, the Enhancing Neuro Imaging Genetics Through Meta-Analysis (ENIGMA) consortium, and the Cohorts for Heart and Aging Research in Genomic Epidemiology (CHARGE) consortium. GWAS data from UKB samples for neuroimaging phenotypes described below were prepared in a similar fashion to UKB GWAS summary data for risk taking phenotypes described above (*see Impulsive Personality Trait GWAS: Genotyping, Imputation, and Quality Control* section above). ENIGMA data (e.g., Grasby et al., 2020; Satizabal et al., 2019) were genotyped using a variety of commercial arrays across the participating studies. Each study sample underwent variant and sample-based QC procedures (e.g., call rate, HWE). Imputation to the 1000 Genomes Project Phase 1 v3 or HRC reference panels was conducted using various validated software packages (see Grasby et al., 2020 and Satizabal et al.,

2019 for additional details). CHARGE data were genotyped to a custom array and imputed to either the European subsamples of the 1000 Genomes Project or HRCr1.1 panels following QC. GWAS summary statistics included morphological and volumetric phenotypes for cortical brain regions and subcortical brain structures. From these sources, three primary sets of imaging phenotypes across the brain were utilized (see Table 1).

**Neuroimaging Phenotypes.** The first set included GWAS summary statistics from UKB obtained using the Oxford Brain Imaging Genetics (BIG40) web server (https://open.win.ox.ac.uk/ukbiobank/big40/) for volumes of 62 cortical regional tissue volume phenotypes (*N* = 31,968; Smith et al., 2021). These include 31 left-hemisphere and 31 right-hemisphere cortical parcellation phenotypes generated via the Desikan-Killiany-Tourville atlas (Klein & Tourville, 2012). The second set included GWAS summary statistics from ENIGMA and UKB obtained by request for cortical surface area and thickness phenotypes controlling for global cortical surface area and thickness (*N* = 33,992; Grasby et al., 2020; (http://enigma.ini.usc.edu/). These include 34 cortical surface area (SA) and 34 cortical thickness (TH) phenotypes generated via the Desikan-Killiany atlas (Desikan et al., 2006). The third set included GWAS summary statistics from CHARGE, ENIGMA, and UKB obtained by request for volumes of seven subcortical structures: the nucleus accumbens, amygdala, caudate nucleus, putamen, globus pallidus, thalamus, and brainstem [including the mesencephalon, pons, and medulla oblongata] (*N* = 24,945 - 30,175; Satizabal et al., 2019; (http://enigma.ini.usc.edu/). CHARGE summary statistics used for this study were from an unrestricted set, and thus, excluded four cohorts restricted from use in research characterized by the study of potentially sensitive behavioral traits (e.g., impulsivity and alcohol use).

### *GenomicSEM Analyses*

Briefly, GenomicSEM is a natural extension of linkage-disequilibrium score regression (LDSC; Bulik-Sullivan, Loh, et al., 2015; Bulik-Sullivan, Finucane, et al., 2015), which calculates genetic correlations between any two traits for which summary statistics are available, provided the samples were drawn from the same ancestral background. Summary statistics are filtered and pre-processed using the *munge* function which retains all HapMap3 SNPs (The International HapMap 3 Consortium, 2010) with MAF > 0.01 outside the major histocompatibility complex region. Using LDSC, GenomicSEM computes a full genetic correlation matrix across the set of traits for which munged GWAS summary statistics are provided and then estimates the model with this correlation matrix using the *lavaan* SEM package (version 0.6-8; Rosseel, 2012) in R. Most of the summary statistics included in the analyses were based on overlapping samples (e.g., 23andMe, UKB); however, this method adjusts for sample overlap by estimating a sampling covariance matrix that indexes the extent to which sampling errors of the estimates are associated (Grotzinger et al., 2019).

Of note, GenomicSEM boosts power relative to GWAS of individual phenotypes even when sample sizes are uneven across phenotypes. However, compared to maximum likelihood estimation, diagonally weighted least squares (DWLS) estimation is more likely to produce a solution that is dominated by the patterns of associations involving the most well-powered traits, thereby producing model estimates that more closely match the pieces of the genetic covariances matrix that are estimated with greater precision (i.e., smaller standard errors typically reflective of larger sample sizes). Importantly, this does not guarantee that model parameters (e.g., factor loadings) will be overly influenced by GWAS with larger sample sizes as several other factors influence the estimation of these parameters including relative correlations between traits, magnitude of differences in relative power of indicator GWAS, degree of sample overlap, and overidentification of models ($df > 0$). Thus, there is a variance-bias tradeoff between DWLS and maximum likelihood estimation here: greater precision of estimates versus decreased bias arising from discrepant sample sizes (Yarkoni & Westfall, 2017). However, in cases of just-identification and high sample overlap, characteristic of single latent factor multivariate GWAS models conducted as part of this study, this tradeoff is expected to be small (Groztinger et al., 2019).

***Multivariate GWAS***

Following examination of structural and stratified GenomicSEM analyses utilizing LD information without measured SNPs, multivariate GWAS analyses were conducted by estimating SNP associations with each latent genetic factor. As a first step in this process, a single set of summary statistics for each model was generated using the *sumstats* function which performs list-wise deletion and standardization across each of the univariate GWAS summary statistics serving as model indicators. Given the list-wise deletion of SNPs missing from any one of the indicators, and thus, variation in the available markers for each latent factor (e.g., sensation seeking vs. others, see Table 1), single latent factor models, rather than two-factor correlated models, were used for multivariate GWAS of dual-systems constructs to optimize power to detect associations for all available SNPs.

Next, the *userGWAS* function was utilized to run the specified structural model in an iterative manner for each measured SNP in the combined set of summary statistics, regressing the latent genetic factor on each SNP individually. In contrast to the structural genomic models, the latent genetic factors were the dependent variables in these analyses, and thus unit loading identification was specified for scaling purposes (i.e., the loading of the GWAS indicator with the greatest correlations with other indicators was set to 1). From these multivariate GWAS analyses, individual SNP associations (unstandardized regression coefficients and standard errors [$b_{SNP}$, $SE_{SNP}$]) with latent factors as well as $\chi^2$ and AIC statistics for each individual SNP regression model were obtained. Individual SNP effects were estimated for the latent genetic factors in each model if they were available in all univariate summary statistics, had a MAF ≥ 0.5%, and were present in the 1000 Genomes Project Phase 3 v5 reference panel.

The effective sample size for each latent factor ($N_{eff}$) was estimated using the approach described by Mallard et al., 2022:

$$N_{eff} = \frac{1}{m} \sum_{MAF=a}^{b} n_j$$

Effective sample size is calculated according to this formula where $n_j$ is the effective sample size of a given SNP for a latent factor, calculated by dividing $(1/SE_{SNP})^2$ by the variance of that SNP based on MAF: 2 × MAF × (1 – MAF). This calculation of $n_j$ produces reasonable estimates for the effective sample size of a given SNP but is prone to errors for SNPs with low MAF. Thus, a limit is set on lower and upper MAF ($a$ = 10% and $b$ = 40%), when estimating the average SNP effective sample ($m$ = number of SNPs) as the total multivariate GWAS effective sample size. Importantly, while it is possible to derive SNP-based heritability estimates ($h_g^2$) from $N_{eff}$, Mallard et al. caution the degree to which these estimates may be interpreted as heritabilities as there is no information about phenotypic variance of the latent genetic factors modeled in GenomicSEM. Thus, $h_g^2$ of latent genetic factors is more accurately referred to as genetic variance and was denoted by $\zeta_g$ to differentiate between these two indices.

In order to assess the extent to which individual SNP effects on each of the indicator phenotypes was not fully mediated by the single latent factor (i.e., common pathway model), follow-up multivariate GWAS models were conducted to calculate $Q_{\text{SNP}}$ tests of heterogeneity. In these secondary models, individual SNP effects are specified such that the influence of each SNP operates through both common and independent pathways (i.e., indicator-specific effects). Nested $\chi^2$ difference tests are then conducted comparing the common pathway versus common pathway plus independent pathways to calculate $Q_{\text{SNP}}$ statistics and associated genome-wide *P*-values for each SNP. In this context, a genome-wide significant (GWS; $P < 5 \times 10^{-8}$) $Q_{\text{SNP}}$ statistic suggests that the independent pathway model (i.e., influence of SNP effects on genetic indicators and latent factor) is a better fit to the data for that SNP than the fully mediated latent factor specification (Grotzinger et al., 2019). Following multivariate GWAS, downstream genetic correlation analyses focused on utilizing SNP associations with these latent genetic factors to differentiate between genetic risk for dual-systems constructs across neuroimaging phenotypes. Thus, SNPs with GWS $Q_{\text{SNP}}$ statistics in each model were removed from model-derived GWAS summary statistics for dual-systems latent factors to reduce heterogeneity in SNP effects on these latent genetic factors prior to downstream genetic correlation analyses.

FUMA (version 1.3.7; Watanabe, Taskesen, van Bochoven, & Posthuma, 2017) was used to define independent GWS SNPs and genomic loci, and to positionally map variants to their nearest gene. Independent GWS SNPs and genomic loci were defined as having an LD $r^2 < .6$ with other GWS SNPs and by a second clumping of independent GWS SNPs by LD $r^2 < .1$ in a 500-kilobase window, respectively.

### References


Bulik-Sullivan, B., Finucane, H. K., Anttila, V., Gusev, A., Day, F. R., Loh, P. R., Duncan, L., Perry, J. R. B., Patterson, N., Robinson, E. B., Daly, M. J., Price, A. L., & Neale, B. M. (2015). An atlas of genetic correlations across human diseases and traits. *Nature Genetics*, *47*(11), 1236–1241. https://doi.org/10.1038/ng.3406

Bulik-Sullivan, B., Loh, P. R., Finucane, H. K., Ripke, S., Yang, J., Patterson, N., Daly, M. J., Price, A. L., Neale, B. M., Corvin, A., Walters, J. T. R., Farh, K. H., Holmans, P. A., Lee, P., Collier, D. A., Huang, H., Pers, T. H., Agartz, I., Agerbo, E., … O'Donovan, M. C. (2015). LD score regression distinguishes confounding from polygenicity in genome-wide association studies. *Nature Genetics*, *47*(3), 291–295. https://doi.org/10.1038/ng.3211

Bycroft, C., Freeman, C., Petkova, D., Band, G., Elliott, L. T., Sharp, K., Motyer, A., Vukcevic, D., Delaneau, O., O'Connell, J., Cortes, A., Welsh, S., Young, A., Effingham, M., McVean, G., Leslie, S., Allen, N., Donnelly, P., & Marchini, J. (2018). The UK Biobank resource with deep phenotyping and genomic data. *Nature*, *562*(7726), 203–209. https://doi.org/10.1038/s41586-018-0579-z

Cloninger C. R., Przybeck T. R., Svrakic D. M., Wetzel R. D. (1994) *The Temperament and Character Inventory (TCI): A guide to its development and use*. St. Louis, MO: Center for Psychobiology of Personality.

Cyders, M. A., Littlefield, A. K., Coffey, S., & Karyadi, K. A. (2014). Examination of a short English version of the UPPS-P Impulsive Behavior Scale. *Addictive Behaviors*, *39*(9), 1372–1376. https://doi.org/10.1016/j.addbeh.2014.02.013

Desikan, R. S., Ségonne, F., Fischl, B., Quinn, B. T., Dickerson, B. C., Blacker, D., Buckner, R. L., Dale, A. M., Maguire, R. P., Hyman, B. T., Albert, M. S., & Killiany, R. J. (2006). An automated labeling system for subdividing the human cerebral cortex on MRI scans into gyral based regions of interest. *NeuroImage*, *31*(3), 968–980. https://doi.org/10.1016/j.neuroimage.2006.01.021

Fuchsberger, C., Abecasis, G. R., & Hinds, D. A. (2015). minimac2: faster genotype imputation. *Bioinformatics*, *31*(5), 782–784. https://doi.org/10.1093/bioinformatics/btu704

Grasby, K. L., Jahanshad, N., Painter, J. N., Colodro-Conde, L., Bralten, J., Hibar, D. P., Lind, P. A., Pizzagalli, F., Ching, C., McMahon, M., Shatokhina, N., Zsembik, L., Thomopoulos, S. I., Zhu, A. H., Strike, L. T., Agartz, I., Alhusaini, S., Almeida, M., Alnæs, D., Amlien, I. K., … Enhancing NeuroImaging Genetics through Meta-Analysis Consortium (ENIGMA)—Genetics working group (2020). The genetic architecture of the human cerebral cortex. *Science, 367*(6484), eaay6690. https://doi.org/10.1126/science.aay6690

Grotzinger, A. D., Rhemtulla, M., de Vlaming, R., Ritchie, S. J., Mallard, T. T., Hill, W. D., Ip, H. F., Marioni, R. E., McIntosh, A. M., Deary, I. J., Koellinger, P. D., Harden, K. P., Nivard, M. G., & Tucker-Drob, E. M. (2019). Genomic structural equation modelling provides insights into the multivariate genetic architecture of complex traits. *Nature Human Behaviour*, *3*(5), 513–525. https://doi.org/10.1038/s41562-019-0566-x

Howie, B., Fuchsberger, C., Stephens, M., Marchini, J., & Abecasis, G. R. (2012). Fast and accurate genotype imputation in genome-wide association studies through pre-phasing. *Nature Genetics*, *44*(8), 955–959. https://doi.org/10.1038/ng.2354

Huang, J., Howie, B., McCarthy, S., Memari, Y., Walter, K., Min, J. L., Danecek, P., Malerba, G., Trabetti, E., Zheng, H. F., UK10K Consortium, Gambaro, G., Richards, J. B., Durbin, R., Timpson, N. J., Marchini, J., & Soranzo, N. (2015). Improved imputation of low-frequency and rare variants using the UK10K haplotype reference panel. *Nature Communications*, *6*, 8111. https://doi.org/10.1038/ncomms9111

Klein, A., & Tourville, J. (2012). 101 labeled brain images and a consistent human cortical labeling protocol. *Frontiers in Neuroscience*, *6*, 171. https://doi.org/10.3389/fnins.2012.00171

Lee, J. J., McGue, M., Iacono, W. G., & Chow, C. C. (2018). The accuracy of LD Score regression as an estimator of confounding and genetic correlations in genome-wide association studies. *Genetic Epidemiology*, *42*(8), 783–795. https://doi.org/10.1002/gepi.22161

Li, H., Handsaker, B., Wysoker, A., Fennell, T., Ruan, J., Homer, N., … 1000 Genome Project Data Processing Subgroup. (2009). The Sequence Alignment/Map format and SAMtools. *Bioinformatics*, *25*(16), 2078–2079. https://doi.org/10.1093/bioinformatics/btp352

Linnér, R. K., Biroli, P., Kong, E., Meddens, S., Wedow, R., Fontana, M. A., Lebreton, M., Tino, S. P., Abdellaoui, A., Hammerschlag, A. R., Nivard, M. G., Okbay, A., Rietveld, C. A., Timshel, P. N., Trzaskowski, M., Vlaming, R., Zünd, C. L., Bao, Y., Buzdugan, L., Caplin, A. H., … Beauchamp, J. P. (2019). Genome-wide association analyses of risk tolerance and risky behaviors in over 1 million individuals identify hundreds of loci and shared genetic influences. *Nature Genetics*, *51*(2), 245–257. https://doi.org/10.1038/s41588-018-0309-3

Mallard, T. T., Linnér, R. K., Grotzinger, A. D., Sanchez-Roige, S., Seidlitz, J., Okbay, A., de Vlaming, R., Meddens, S. F. W., Palmer, A. A., Davis, L. K., Tucker-Drob, E. M., Kendler, K. S., Keller, M. C., Koellinger, P. D. & Harden, K. P. (2022). Multivariate GWAS of psychiatric disorders and their cardinal symptoms reveal two dimensions of cross-cutting genetic liabilities. *Cell Genomics*, *2*(6), 100140. https://doi.org/10.1016/j.xgen.2022.100140

McCarthy, S., Das, S., Kretzschmar, W., Delaneau, O., Wood, A. R., Teumer, A., Kang, H. M., Fuchsberger, C., Danecek, P., Sharp, K., Luo, Y., Sidore, C., Kwong, A., Timpson, N., Koskinen, S., Vrieze, S., Scott, L. J., Zhang, H., Mahajan, A., Veldink, J., … Haplotype Reference Consortium (2016). A reference panel of 64,976 haplotypes for genotype imputation. *Nature Genetics*, *48*(10), 1279–1283. https://doi.org/10.1038/ng.3643

Miettunen, J., Kantojärvi, L., Ekelund, J., Veijola, J., Karvonen, J.T., Peltonen, L. Järvelin, M. R., Freimer, N., Lichtermann, D., & Joukamaa, M. (2004). A large population cohort provides normative data for investigation of temperament. *Acta Psychiatrica Scandinavica, 110*, 150–157. https://doi.org/10.1111/j.1600-0047.2004.00344.x

Patton, J., Stanford, M., & S. Barratt, E. (1995). Factor structure of the Barratt Impulsiveness Scale. *Journal of Clinical Psychology*, *51*, 768–774. https://doi.org/10.1002/1097-4679(199511)51:63.0.CO;2-1

Rosseel, Y. (2012). lavaan: An R package for structural equation modeling. *Journal of Statistical Software*, *48*, 1-36. https://doi.org/10.18637/jss.v048.i02

Rüeger, S., McDaid, A., & Kutalik, Z. (2018). Evaluation and application of summary statistic imputation to discover new height-associated loci. *PLoS Genetics*, *14*(5), e1007371. https://doi.org/10.1371/journal.pgen.1007371

Sanchez-Roige, S., Fontanillas, P., Elson, S. L., Gray, J. C., De Wit, H., MacKillop, J., & Palmer, A. A. (2019). Genome-wide association studies of impulsive personality traits (BIS-11 and UPPS-P) and drug experimentation in up to 22,861 adult research participants identify loci in the CACNA1I and CADM2 genes. *Journal of Neuroscience*, *39*(13), 2562–2572. https://doi.org/10.1523/JNEUROSCI.2662-18.2019

Satizabal, C. L., Adams, H., Hibar, D. P., White, C. C., Knol, M. J., Stein, J. L., Scholz, M., Sargurupremraj, M., Jahanshad, N., Roshchupkin, G. V., Smith, A. V., Bis, J. C., Jian, X., Luciano, M., Hofer, E., Teumer, A., van der Lee, S. J., Yang, J., Yanek, L. R., Lee, T. V., … Ikram, M. A. (2019). Genetic architecture of subcortical brain structures in 38,851 individuals. *Nature Genetics*, *51*(11), 1624–1636. https://doi.org/10.1038/s41588-019-0511-y

Service, S. K., Verweij, K. J. H., Lahti, J., Congdon, E., Ekelund, J., Hintsanen, M., Räikkönen, K., Lehtimäki, T., Kähönen, M., Widen, E., Taanila, A., Veijola, J., Heath, A. C., Madden, P. A. F., Montgomery, G. W., Sabatti, C., Järvelin, M. R., Palotie, A., Raitakari, O., … Freimer, N. B. (2012). A genome-wide meta-analysis of association studies of Cloninger's Temperament Scales. *Translational Psychiatry*, *2*, 1–9. https://doi.org/10.1038/tp.2012.37

Smith, S. M., Douaud, G., Chen, W., Hanayik, T., Alfaro-Almagro, F., Sharp, K., & Elliott, L. T. (2021). An expanded set of genome-wide association studies of brain imaging phenotypes in UK Biobank. *Nature Neuroscience*, *24*(5), 737–745. https://doi.org/10.1038/s41593-021-00826-4

The 1000 Genomes Project Consortium, Abecasis, G. R., Altshuler, D., Auton, A., Brooks, L. D., Durbin, R. M., Gibbs, R. A., Hurles, M. E., & McVean, G. A. (2010). A map of human genome variation from population-scale sequencing. *Nature*, *467*(7319), 1061–1073. https://doi.org/10.1038/nature09534

The 1000 Genomes Project Consortium. (2015). A global reference for human genetic variation. *Nature*, *526*(7571), 68–74. https://doi.org/10.1038/nature15393

The International HapMap 3 Consortium, Altshuler, D. M., Gibbs, R. A., Peltonen, L., Altshuler, D. M., Gibbs, R. A., Peltonen, L., Dermitzakis, E., Schaffner, S. F., Yu, F., Peltonen, L., Dermitzakis, E., Bonnen, P. E., Altshuler, D. M., Gibbs, R. A., de Bakker, P. I., Deloukas, P., Gabriel, S. B., Gwilliam, R., Hunt, S., … McEwen, J. E. (2010). Integrating common and rare genetic variation in diverse human populations. *Nature*, *467*(7311), 52–58. https://doi.org/10.1038/nature09298

The UK10K Consortium, Walter, K., Min, J. L., Huang, J., Crooks, L., Memari, Y., McCarthy, S., Perry, J. R., Xu, C., Futema, M., Lawson, D., Iotchkova, V., Schiffels, S., Hendricks, A. E., Danecek, P., Li, R., Floyd, J., Wain, L. V., Barroso, I., Humphries, S. E., … Soranzo, N. (2015). The UK10K project identifies rare variants in health and disease. *Nature*, *526*(7571), 82–90. https://doi.org/10.1038/nature14962

Watanabe, K., Taskesen, E., van Bochoven, A., & Posthuma, D. (2017). Functional mapping and annotation of genetic associations with FUMA. *Nature Communications*, *8*(1), 1826. https://doi.org/10.1038/s41467-017-01261-5

Willer, C. J., Li, Y., & Abecasis, G. R. (2010). METAL: fast and efficient meta-analysis of genomewide association scans. *Bioinformatics*, *26*(17), 2190–2191. https://doi.org/10.1093/bioinformatics/btq340

Yarkoni, T., & Westfall, J. (2017). Choosing prediction over explanation in psychology: Lessons from machine learning. *Perspectives on Psychological Science*, *12*(6), 1100–1122. https://doi.org/10.1177/1745691617693393
