## Supplementary Figures for "Dual-systems models of the genetic architecture of impulsive personality traits: Neurogenetic evidence of distinct but related factors"

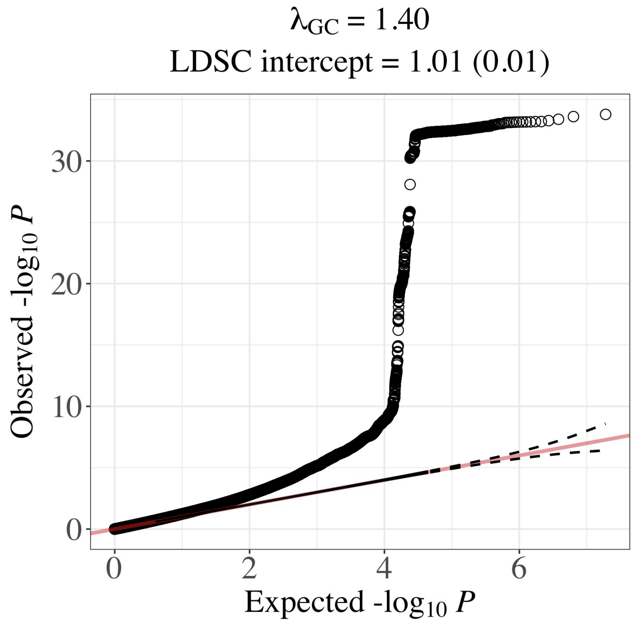


**Supplementary Figure 1.** Q-Q plot for GenomicSEM indicator GWAS meta-analyses of 23andMe + Linnér et al. (2019) risk taking. These results were not adjusted for genomic control inflation factors (λGC).

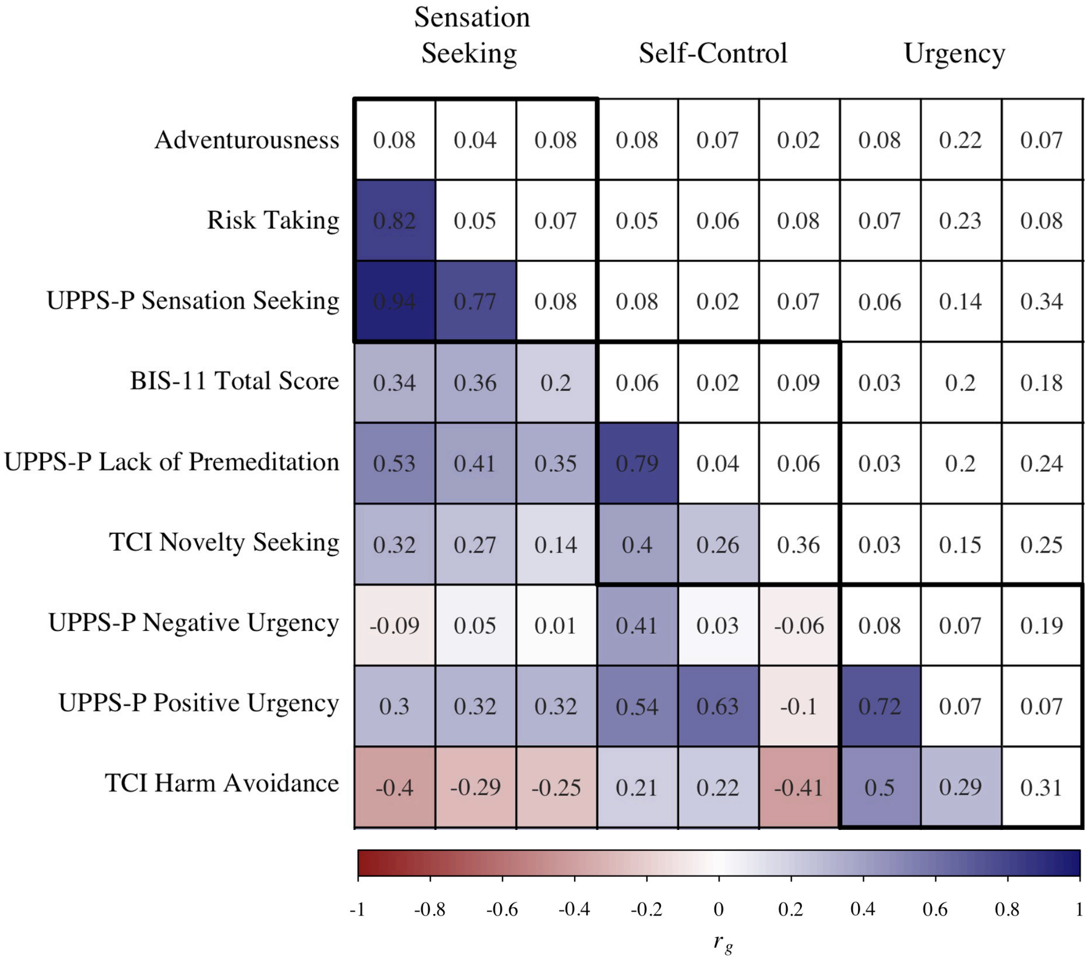


**Supplementary Figure 2. Genetic correlations among factor indicators in GenomicSEM single factor models.** The lower and upper triangles display pairwise LDSC genetic correlation ($r_g$) estimates and their *SE*, respectively, for indicator GWAS in final models (see Supplementary Tables 1 and 2). Diagonal displays observed-scale SNP heritability estimates ($h_g^2$). Indicator GWAS are grouped by latent factors (top of matrix) and intercorrelations and *SE*s within each factor are surrounded by black squares. Note, “self-control” denotes (lack of) self-control factor.

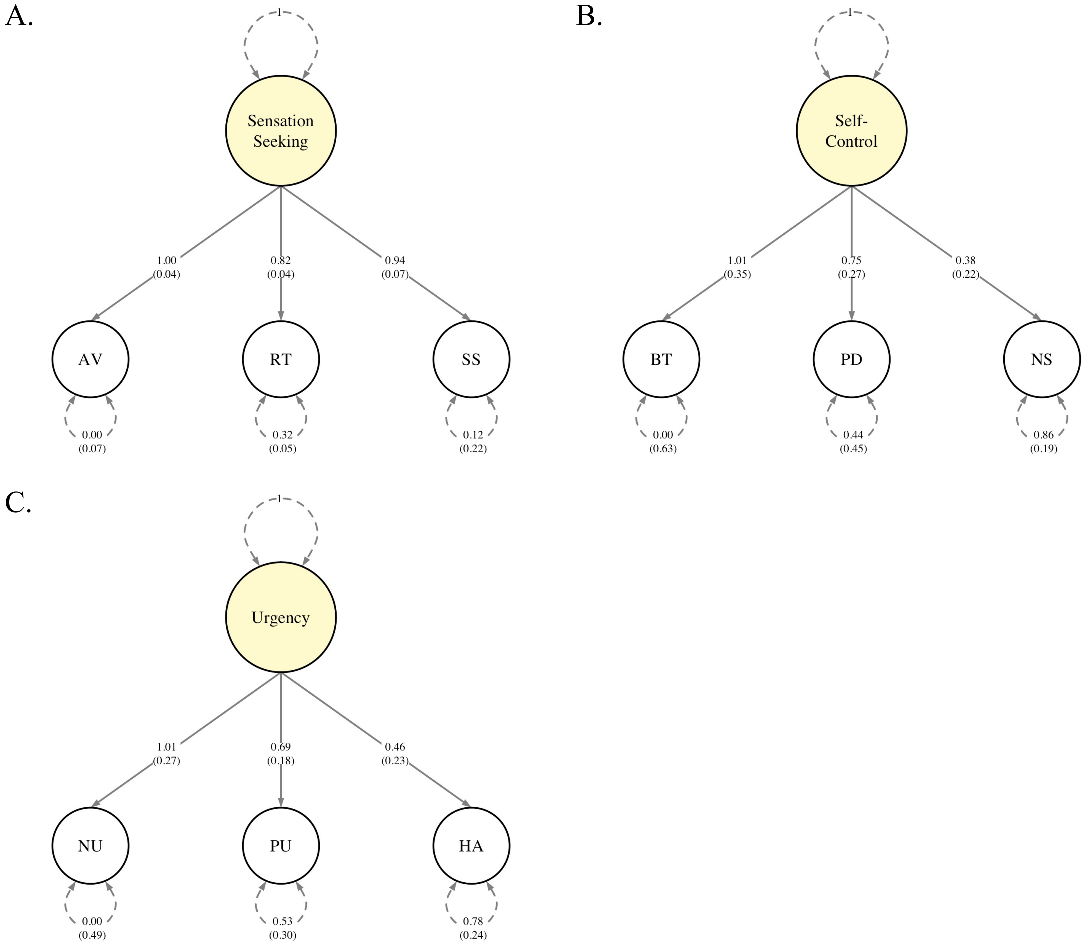


**Supplementary Figure 3. Final path diagram of the single factor dual-systems constructs estimated with GenomicSEM. (A.)** Sensation seeking factor. **(B.)** (Lack of) self-control factor. **(C.)** Urgency factor. Presented parameters are standardized and *SE* are shown in paratheses. Variances are shown as dashed lines and factor loadings shown as solid lines (Supplementary Table 5). Note, "self-control" denotes (lack of) self-control factor. AV = Adventurousness; RT = Risk Taking; SS = UPPS-P Sensation Seeking; BT = BIS-11 Total Score; PD = UPPS-P Lack of Premeditation; NS = TCI Novelty Seeking; NU = UPPS-P Negative Urgency; PU = UPPS-P Positive Urgency; HA = TCI Harm Avoidance

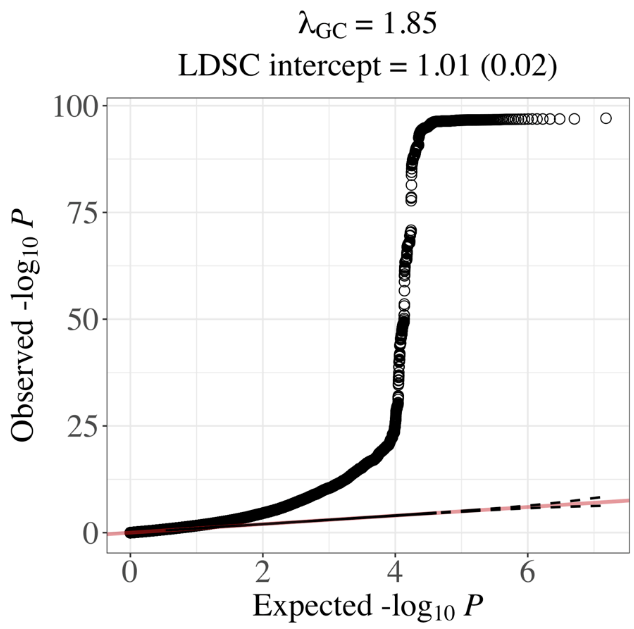


**Supplementary Figure 4.** Q-Q plot for GenomicSEM multivariate GWAS of sensation seeking factor. These results were not adjusted for genomic control inflation factors (λGC).

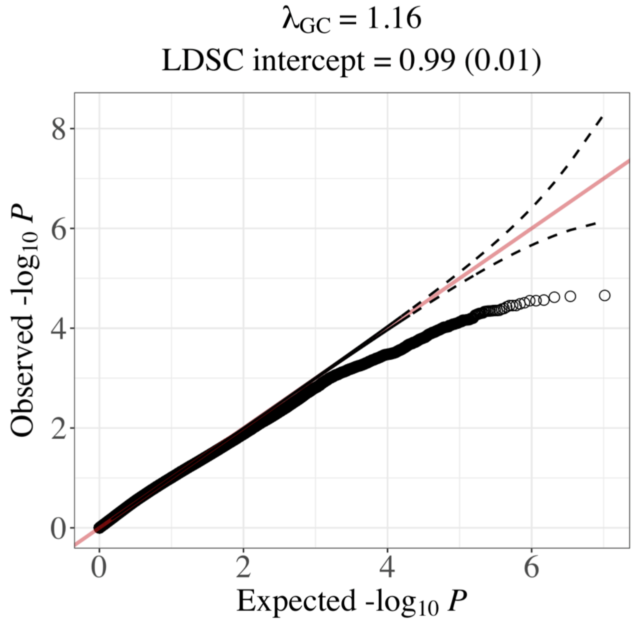


**Supplementary Figure 5.** Q-Q plot for GenomicSEM multivariate GWAS of (lack of) self-control factor. These results were not adjusted for genomic control inflation factors (λGC).

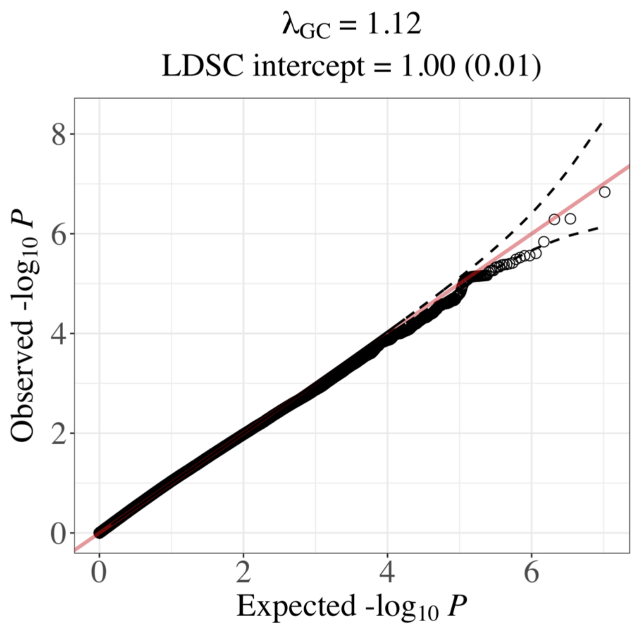


**Supplementary Figure 6.** Q-Q plot for GenomicSEM multivariate GWAS of urgency factor. These results were not adjusted for genomic control inflation factors (λGC).
